# A Systematic Review and Meta-Analysis of Digital Application use in Clinical Research in Pain Medicine

**DOI:** 10.1101/2022.03.02.22271773

**Authors:** Ashish Shetty, Gayathri Delanerolle, Yutian Zeng, Jian Qing Shi, Rawan Ebrahim, Joanna Pang, Dharani Hapangama, Martin Sillem, Suchith Shetty, Balakrishnan Shetty, Martin Hirsch, Vanessa Raymont, Kingshuk Majumder, Sam Chong, William Goodison, Rebecca O’Hara, Louise Hull, Nicola Pluchino, Naresh Shetty, Sohier Elneil, Tacson Fernandez, Peter Phiri, Robert Brownstone

## Abstract

**Importance:** Pain is a silent global epidemic impacting approximately a third of the population. Pharmacological and surgical interventions are primary modes of treatment. Cognitive/behavioural management approaches and interventional pain management strategies are approaches that have been used to assist with the management of chronic pain. Accurate data collection and reporting treatment outcomes are vital to addressing the challenges faced. In light of this, we conducted a systematic evaluation of the current digital application landscape within chronic pain medicine.

**Objective:** The primary objective was to consider the prevalence of digital application usage for chronic pain management. These digital applications included mobile apps, web apps, and chatbots.

**Data Sources:** We conducted searches on PubMed and ScienceDirect for studies that were published between 1st January 1990 and 1st January 2021.

**Study Selection:** Our review included studies that involved the use of digital applications for chronic pain conditions. There were no restrictions on the country in which the study was conducted. Only studies that were peer-reviewed and published in English were included. Four reviewers had assessed the eligibility of each study against the inclusion/exclusion criteria. Out of the 84 studies that were initially identified, 38 were included in the systematic review.

**Data Extraction and Synthesis:** The AMSTAR guidelines were used to assess data quality. This assessment was carried out by 3 reviewers. The data were pooled using a random-effects model.

**Main Outcome(s) and Measure(s):** Before data collection began, the primary outcome was to report on the prevalence of digital application usage for chronic pain conditions. We also recorded the type of digital application studied (e.g. mobile application, web application) and, where the data was available, the prevalence of pain intensity, pain inferences, depression, anxiety, and fatigue.

**Results:** 38 studies were included in the systematic review and 22 studies were included in the meta-analysis.

The digital interventions were categorised to web and mobile applications and chatbots, with pooled prevalence of 0.22 (95% CI −0.16, 0.60), 0.30 (95% CI 0.00, 0.60) and −0.02 (95% CI −0.47, 0.42) respectively. Pooled standard mean differences for symptomatologies of pain intensity, depression, and anxiety symptoms were 0.25 (95% CI 0.03, 0.46), 0.30 (95% CI 0.17, 0.43) and 0.37 (95% CI 0.05, 0.69) respectively.

A sub-group analysis was conducted on pain intensity due to the heterogeneity of the results (I^2^=82.86%; p=0.02). After stratifying by country, we found that digital applications were more likely to be effective in some countries (e.g. USA, China) than others (e.g. Ireland, Norway).

**Conclusions and Relevance:** The use of digital applications in improving pain-related symptoms shows promise, but further clinical studies would be needed to develop more robust applications.

## Introduction

High-quality research data generated by scientifically robust study designs, improved use of clinical data, and the development of cost-effective healthcare models can change how medicine is practiced in the modern world. Digital medicine (DM), wherein multimodal and multidimensional digital tools are used to intervene in accessing and providing healthcare, is now a fundamental part of these drivers of change.

Despite relative growth profoundly impacting gross economic improvement, ‘*bench to bedside*’ pathways still take considerable time (). Equally robust research evaluations have not kept pace with a growing global population, although, the intellectual and healthcare evolution has modernised clinical practice by way of clinical research. Existing clinical evidence and incorporation of information technology has led to more prominent use of DM. A fundamental aspect of DM is to improve and promote evidence-based medicine (EBM) and/or evidence-based practices (EBP) within clinical and healthcare frameworks, underpinned by data science and technologies.

The field of pain medicine in adults is a particularly challenging area of clinical practice for many reasons, including subjectivity associated with patient-reported outcomes and management of symptomatology with limited information on pathophysiology (). Considering this uncertainty, attempts by clinicians to categorise pain and decide on treatment interventions (Supplementary Table 1), could benefit from the concepts of DM and its associates of EBM and EBP. Pain is often the commonest symptom that patients present with in outpatient clinics. The need for individualised care based on generalisable research is complicated by wide variables, subjective nature, and inherent bias which provide a unique set of challenges for a simple protocol to work. The use of cognitive technology such as artificial intelligence, in delivering personalised care, based on available evidence, is therefore an attractive proposition for pain medicine.

Pain medicine has been identified as a specialty that would vastly benefit from the personalisation of care.^1^ A current example of this need is the variable efficacy of pharmacotherapy in relieving chronic pain. Opioids, for instance, have been routinely used to treat chronic pain syndromes, despite only modest evidence for their use.^2^ This has the potential for significant harm in patients where it has been used inappropriately and may have influenced factors that led to the Opioid Crisis globally, especially so in the USA and UK. Traditional pain evaluation methods are vulnerable to recall error and bias as they rely on retrospective reporting of pain variations.^3^ Pain perception combined with measuring functional changes and physiological parameters affected by pain are important secondary outcome data to assess efficacy. Methods demanding frequent, repeated pain evaluation and pain-associated features are required to formulate chronic pain management strategies.^4,5^ This approach was previously hindered both by the resources required for such vast data collection, and the complexity of the statistical analysis required to interpret the resulting datasets.^6^

To advance DM concepts and their use in pain medicine research, it is imperative to assess the global regulatory sphere. Over the last decade, a plethora of legislations and regulatory guidelines around DM have been developed by the World Health Organisation (WHO),^7^ Medicines and Healthcare products Regulatory Agency (MHRA),^8^ Food and Drug Administration (FDA)^9^ and National Institute for Health and Care Excellence (NICE)^10^ (Supplementary Table 2). However, there are complexities around evaluating Artificial Intelligence (AI)-based applications that fall under the category of DM. This includes those using algorithms based on machine learning (ML) models that may be categorised as a medical device. Furthermore, development of AI applications requires documentary evidence that the planning, designing, and development phases meet the globally accepted Internationally Organisation for Standardisation (ISO) standards. In order to achieve ISO standards, a high proficiency of conformances should be maintained by the research group responsible for developing the intervention that could be mass produced. As part of this standardisation process, the intervention may undergo several non-conformity assessments as well as vigorous testing and validation prior to being deployed.

The regulatory and standards required for novel innovations are also dependent on the disease classification. The current classification of chronic and acute pain conditions (Figure 1 and Figure 2 respectively) employs the guidelines published by the International Association for the Study of Pain. Clinicians evaluating both chronic and acute chronic pain are considering changes to guidelines to provide better diagnoses and improve outcomes for patients. Advancements in the understanding of the pathophysiology of acute and chronic pain have resulted in effective pharmacological approaches to sub-populations of patients.

**Figure 1.**
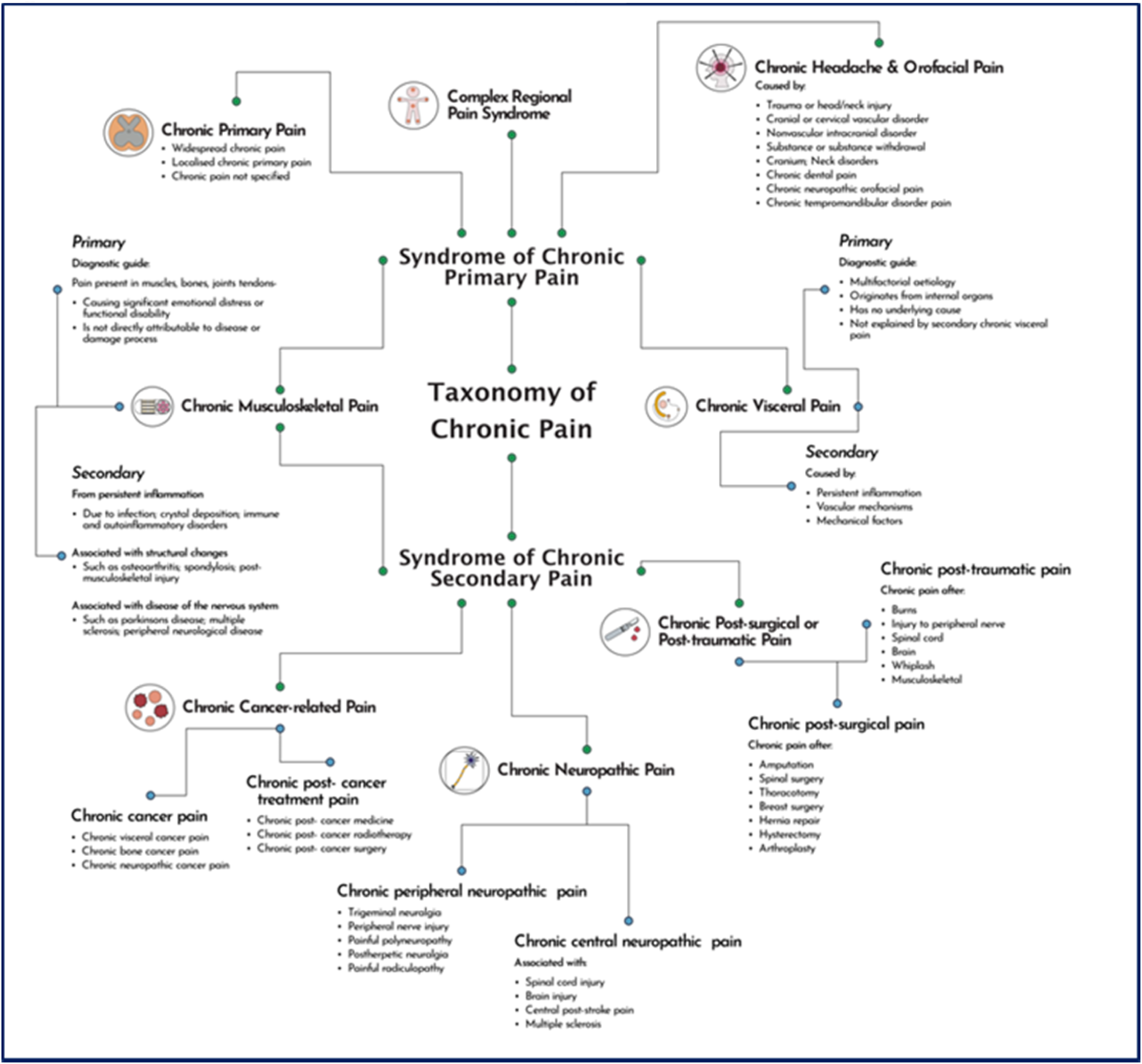
Chronic Pain Classification Tree (CPCT)

**Figure 2.**
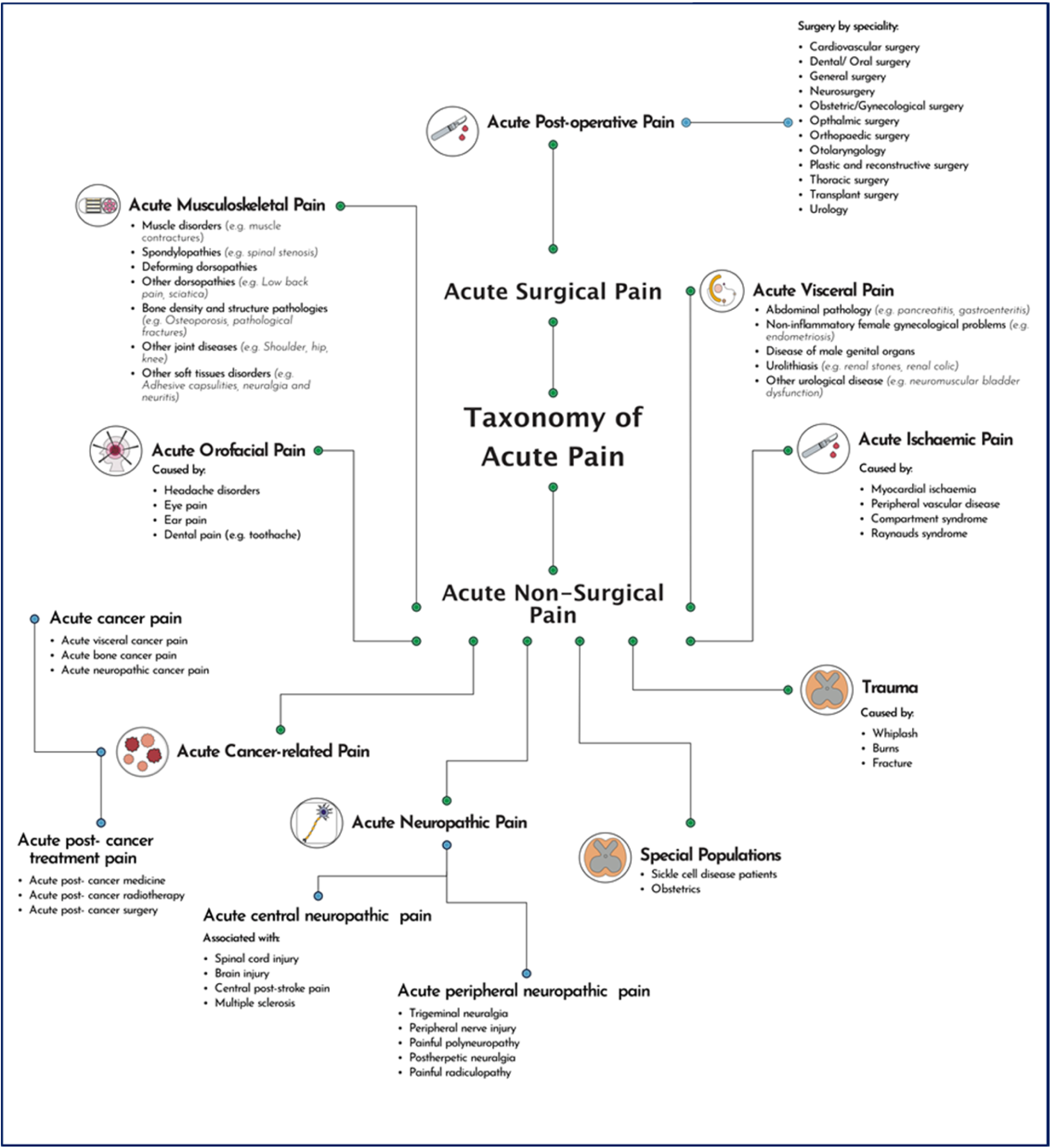
Acute Pain Classification Tree

A critical step of DM is the development of digital tools using large sets of datasets and aggregated data to create novel paradigms of care. This is also referred to as evidence-based digital medicine which uses EBM concepts. To disperse these paradigms, computer programming, utility and broad access of applications are vital. The development of smartphone applications is key to deliver the DM phenomenon to facilitate communication and engagement between clinicians and patients. A key element would be to personalise both treatments and applications using sensors and programming capabilities that would support significant benefits as summarised in Supplementary Table 3.

Evaluating the current DM landscape is equally important as developing novel applications. The accessibility of smartphones has given rise to multiple pilots of app-based longitudinal assessment programmes for chronic pain, which have shown promising early results.^11,12^ Furthermore, the use of validated lifestyle devices such as the FitBit® as monitoring adjuncts could be combined with questionnaires and activity programmes to allow regular functional reassessment among chronic pain patients.^13^

Therefore, the primary aims of this study were to: (1) identify and report the current prevalence of DM application in pain medicine; (2) identify and report the current DM application use within pain medicine. To achieve this, we aimed to explore the prevalence of these types of assessments’ use and deployment of these using DM applications.

## Materials and methods

An evidence synthesis methodology was developed for the purpose of this study, with a systematic review protocol published on PROSPERO (CRD42021248232). The Preferred Reporting Items for Systematic Reviews and Meta-analyses (PRISMA) was used to report findings.

### Search strategy and study selection

PubMed and ScienceDirect were used to identify relevant studies that were peer-reviewed and published in English between the 1st of January 1990 and 1st of January 2021. Search terms used included *Chronic Pain, Pain Clinical Trials, Pain medicine, Pain medicine clinical research* and *Digital Clinical Trials*. All studies using DM applications for chronic pain conditions were included. Only studies that were peer-reviewed and published in English were included. Suitable publications were selected using the PICO (Population/Participants, Intervention(s), Comparison, Outcome) strategy. An independent reviewer screened studies included within the study by reading the full text. Initial title and abstracts for identified articles were screened by 4 investigators. Inclusion and exclusion criteria were assessed against each study. This was followed with the screening of the full study article independently by 2 investigators and included into the final data pool.

### Data extraction and synthesis

The data extraction process involved reading titles and abstracts followed by the application of the refinement protocol where the full text was reviewed and subsequently verified. Key study details such as study title, citation details, methods, findings, limitations, characteristics of the study and conclusions were extracted. Differing opinions were resolved by review and discussion between the lead authors. The authors remained unblinded regarding the publisher details. A full methodological description is demonstrated within the supplementary document (Figure 3).

**Figure 3.**
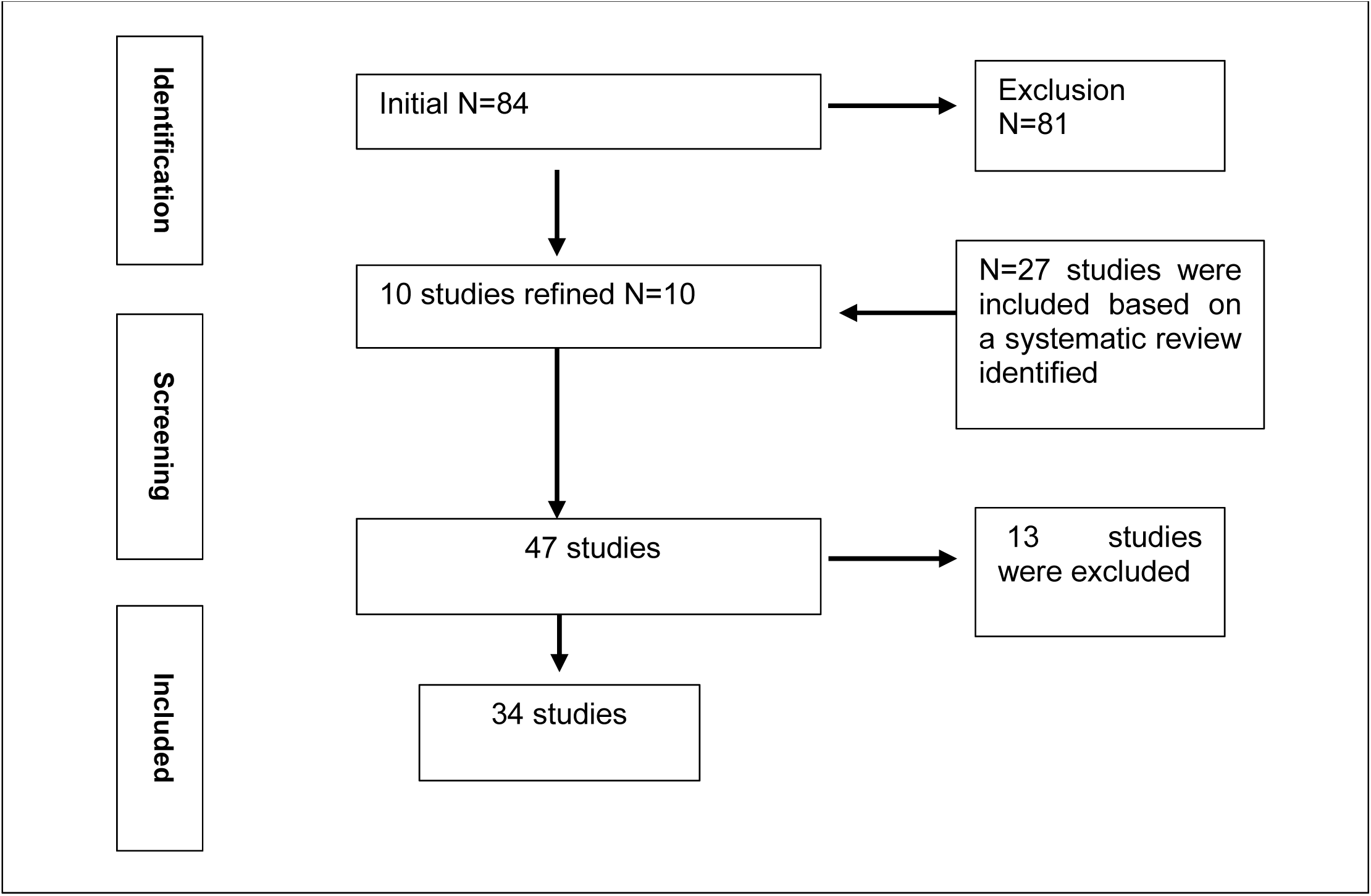
Representation of the PRISMA Flowchart

### Data analysis

As all studies reported the mean and SD at several time points, a mathematical model was formulated as demonstrated in Supplementary Figure 1.

BPI, NRS, PCP-S and pain evaluation questionnaires were used to assess pain intensity and pain interference; HADS, CES-D, BDI, PHQ-9, DASS, GAD-7 and STAI were used to assess depression and anxiety; FSS and MOS sleep scale were used to assess fatigue and sleep. All studies reported the mean and SD of the questionnaires across several timepoints, at baseline and follow-up. The baseline questionnaire score was subtracted from the follow-up questionnaire score to standardize the data and remove the initial effect. Score changes between these two time points reflect the treatment effect. 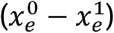 represented the change in the questionnaire scores between baseline (^0^) and follow-up (^1^) in the treatment group, which also indicated an improvement of treatments, and 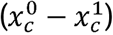 represented the change in the questionnaire scores between baseline (^0^) and follow-up (^1^) in the control group.

Therefore, 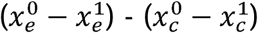 showed the mean difference (MD) of the change of score between the two groups, which is the outcome of focus. If 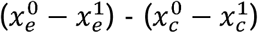 is positive, it indicates the treatment was beneficial for patients in improving symptoms of pain. However, if 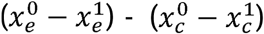 is negative, it indicates the treatment had no effect on improving pain.

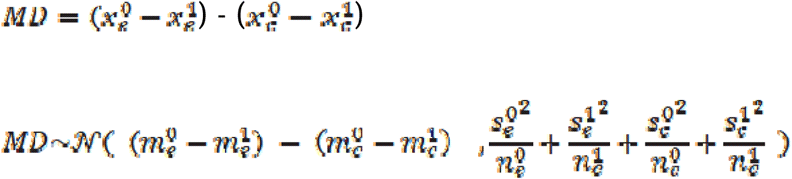

The scales of the questionnaires were different, therefore standardized mean differences (SMD) were used to illustrate the change in the mean score of the treatment group versus the control group from baseline to follow-up. The traditional form of SMD was

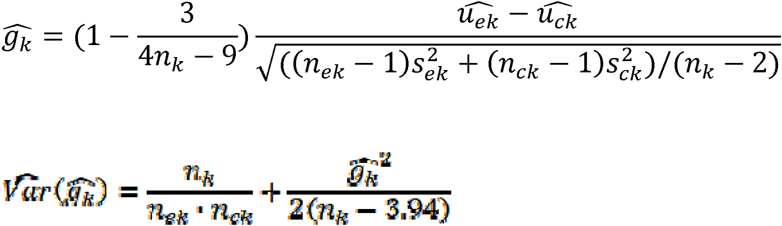

where 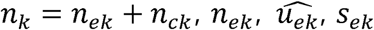 are the number, mean and standard variation of treatment group. 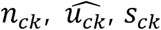 are the number, mean and standard variation of the control group. The 95% confidence interval (CI) was obtained by

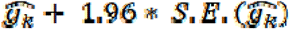

where 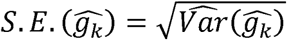.

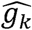 was transformed according to the traditional form, and 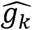 and 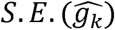 were calculated for each study, with a random effect model used to pool the estimators. Funnel plot graphs demonstrated the publication bias. Subgroup analysis and I^2^ were used to explain heterogeneity and Egger’s test was used to detect publication bias. All procedures were finished with STATA 16.1.

### Risk of bias

The risk of bias (RoB) table (below) has been used to demonstrate the risk of bias within the randomised controlled trials used in the systematic review and meta-analysis. The RoB is reflective of a fixed set of biases within domains of study design, conduct and reporting. This combined with the quality check allows the findings of the study to be scientifically justified, and clinically viable.

AMSTAR was used also to assess methodological quality, where the total scores range from 0-11 (see Figure 4, below). An article would be considered as good quality with a score of 8-11, moderate 4-7 and low 0-3.

**Figure 4.**
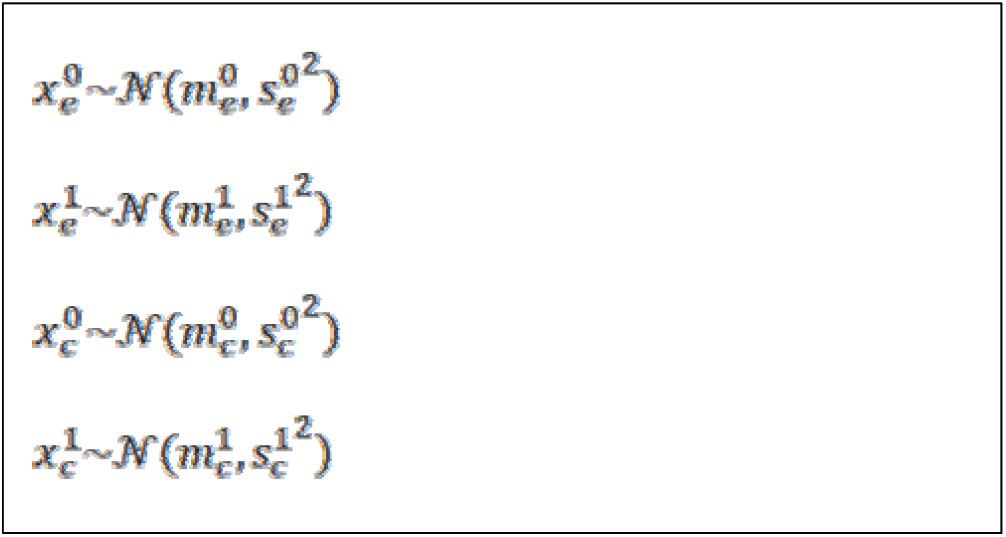
Mathematical model for data analysis

### Outcomes

Outcomes of this study were reported via the meta-analysis which was based on the availability of statistics reported by the systematically included studies. The following are the outcomes of this study:

- Prevalence of DM applications, including categories
- Prevalence of chronic pain conditions using DM applications for self-reporting purposes
- Prevalence of pain outcomes of depression, anxiety, pain inferences, and fatigue and sleep problems
- Clinical significance of the prevalence data
- Research significance of the prevalence data
- Critical interpretation of the identified data
- Common themes identified within the prevalence data

## Results

The search yielded 84 publications, with 38^11,16-52^ included as part of the systematic review (Table 1). Of the 38 studies, 7 were cross-sectional and lacked a control group. Eight studies comprised of a control and treatment group, although they either lacked statistical information completely or inconsistencies were identified that were associated with the mean and SD at baseline and beta coefficients at follow-up timepoints. Therefore, 16^25,28,29,31,33,35-40,43,46,50-52^ were excluded and 22^11,16-24,26,27,30,32,34,41,42,44,45,47-49^ were included into the final meta-analysis (Table 2).

**Table 1.**
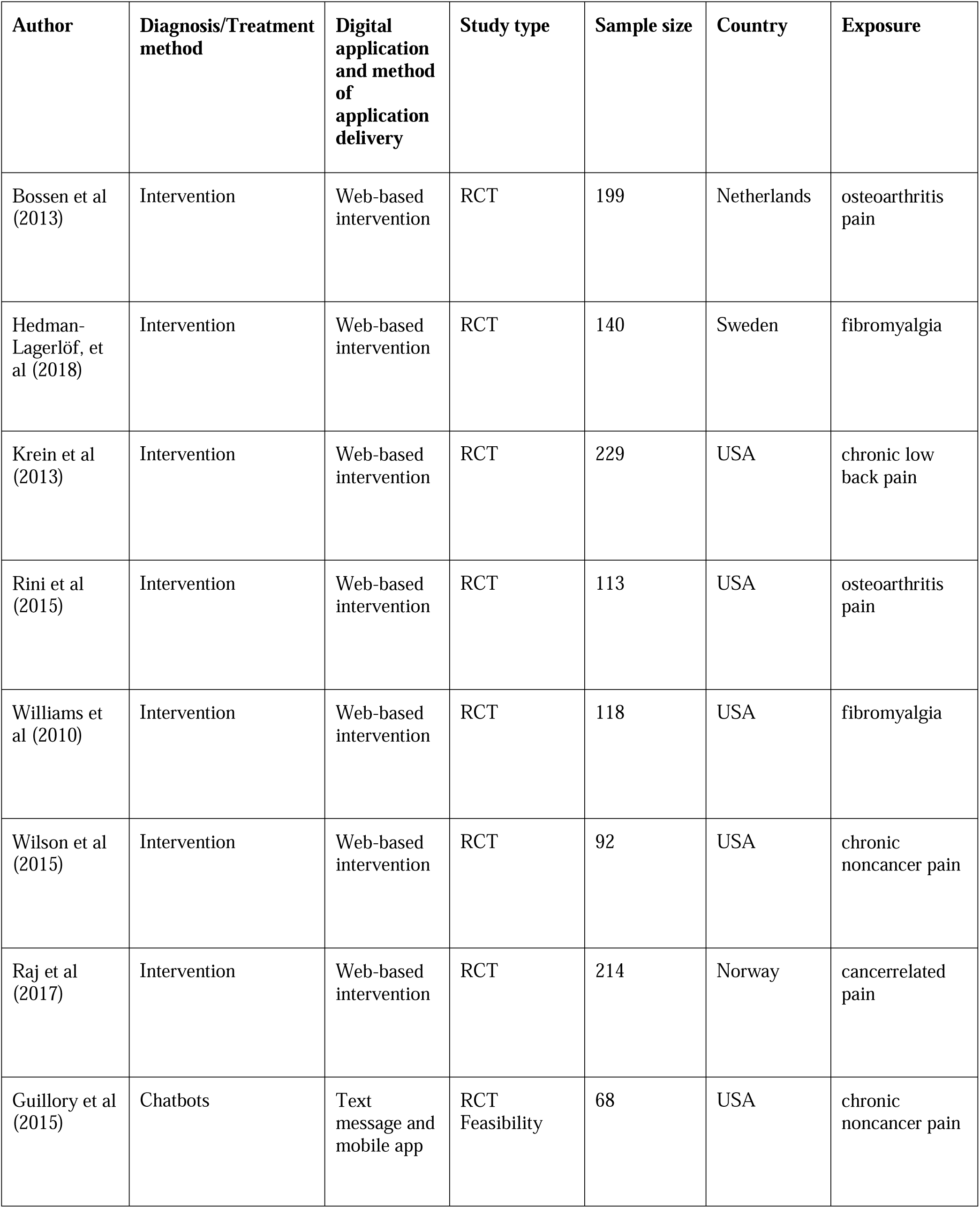
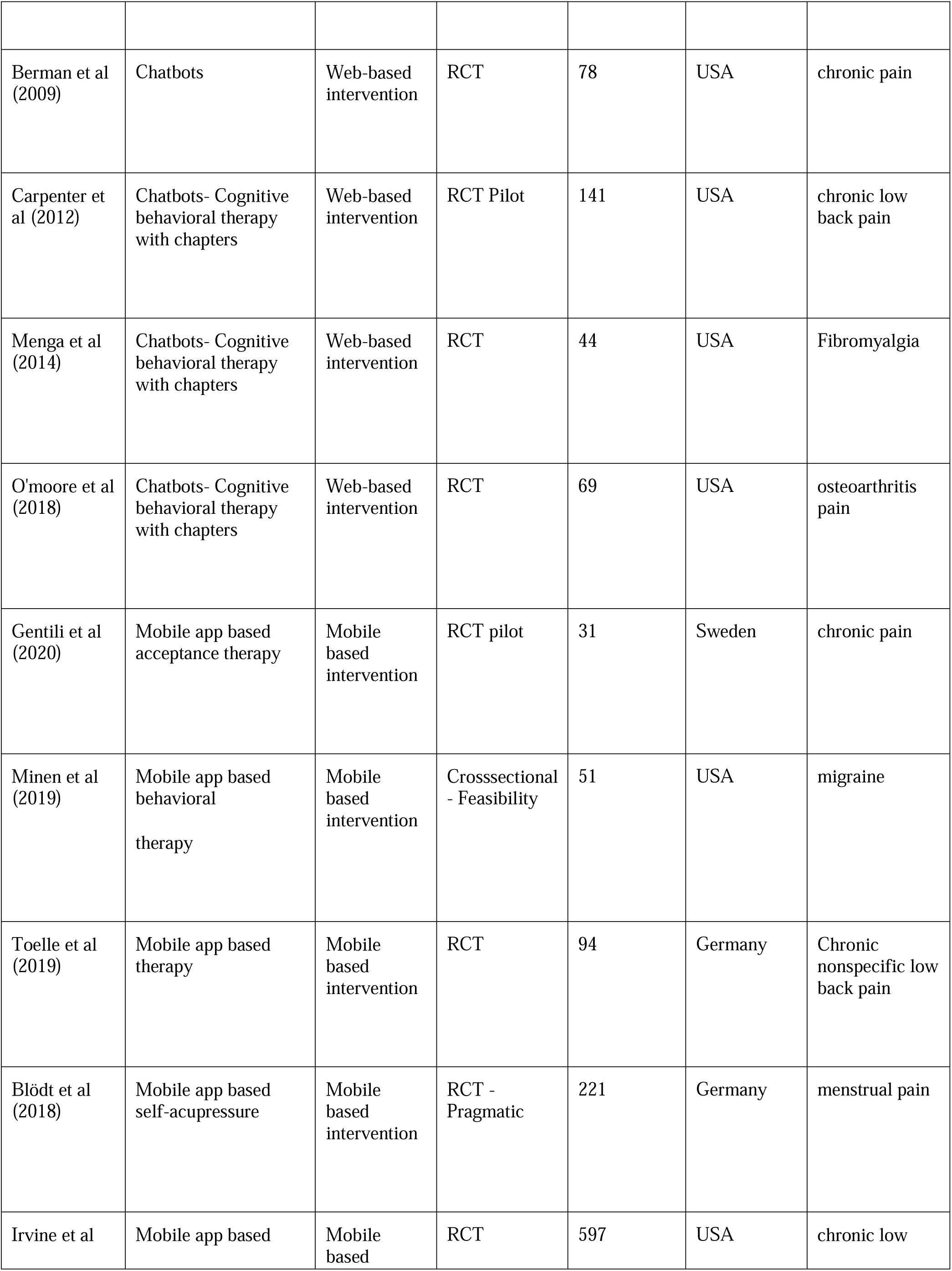
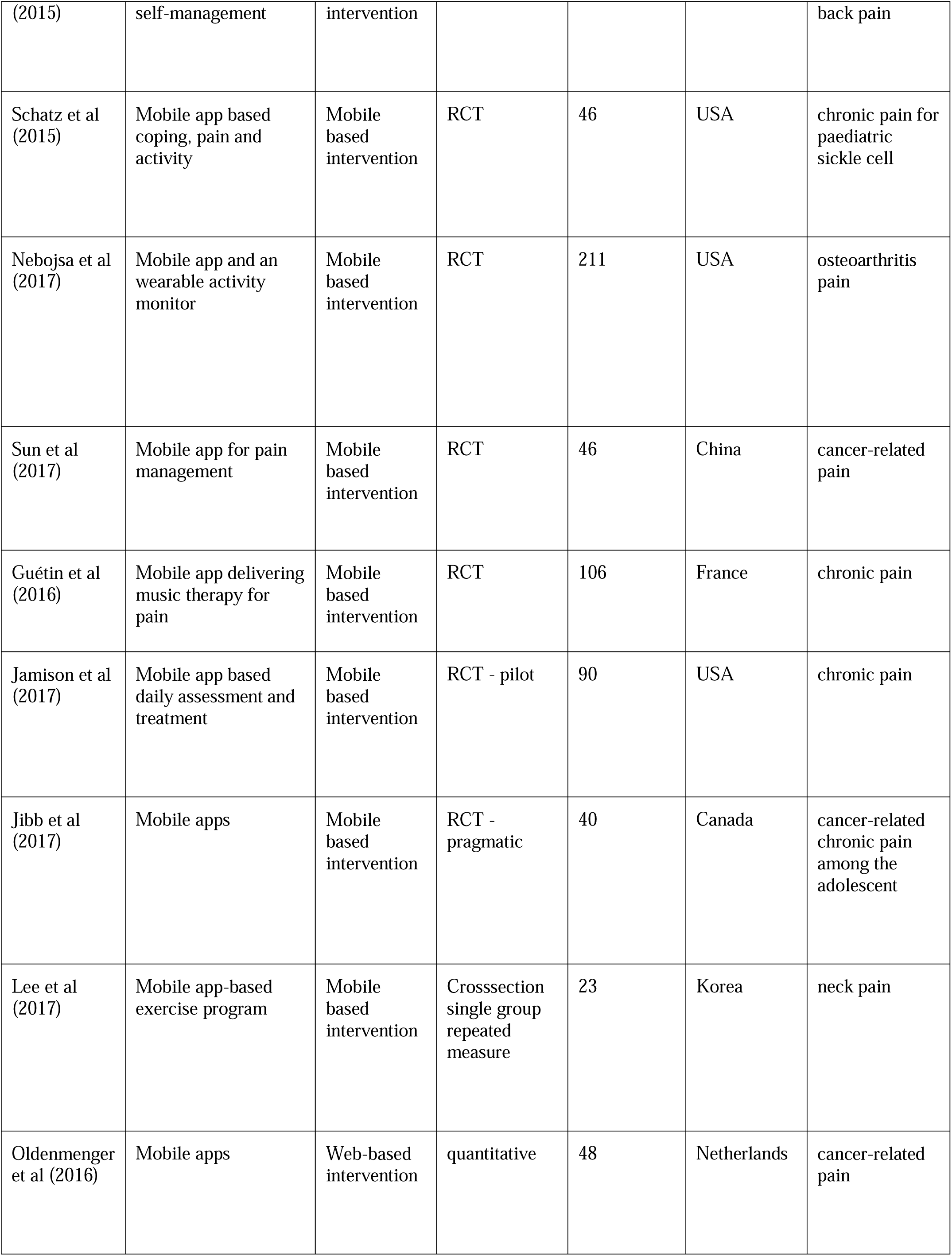
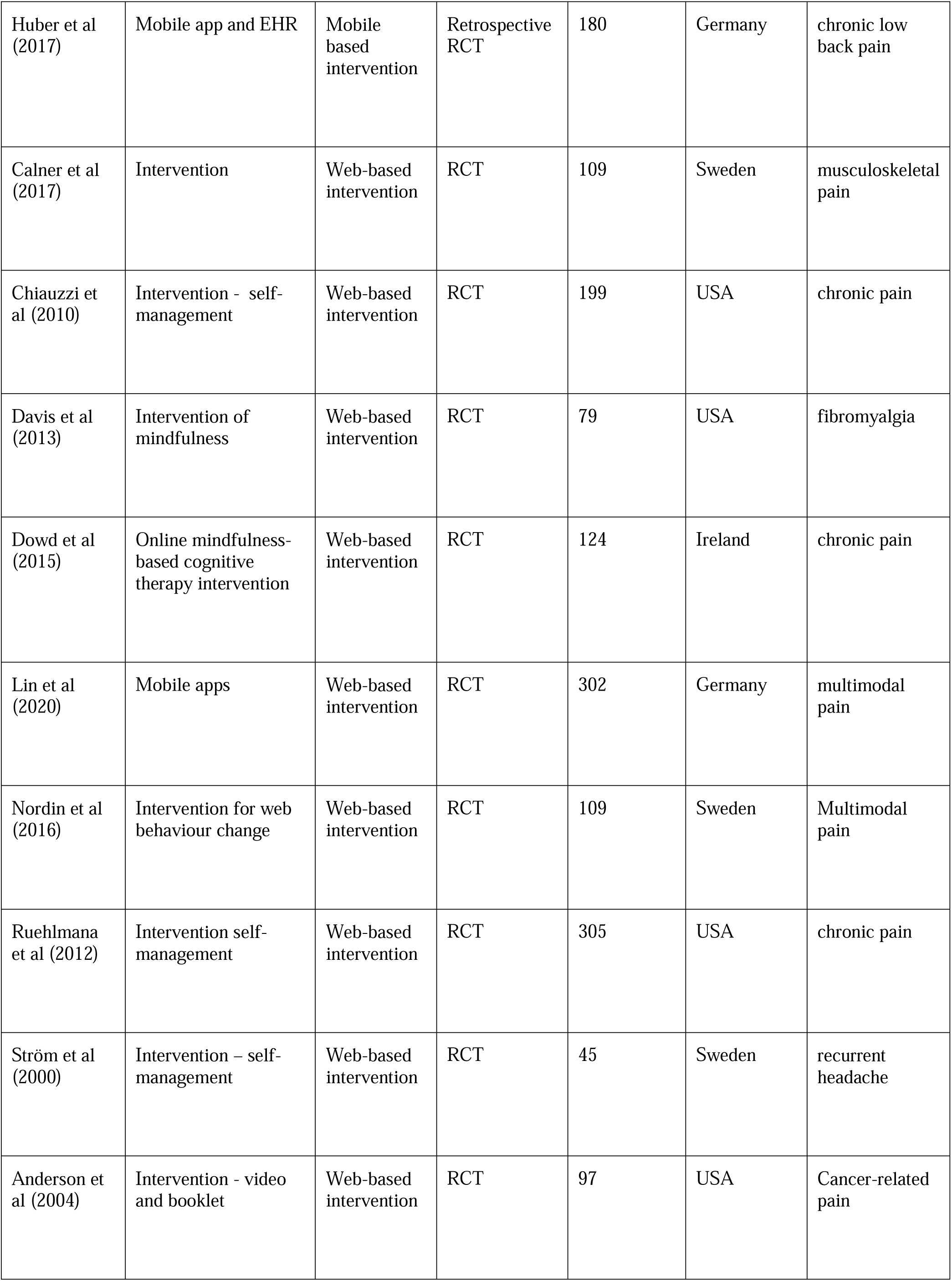
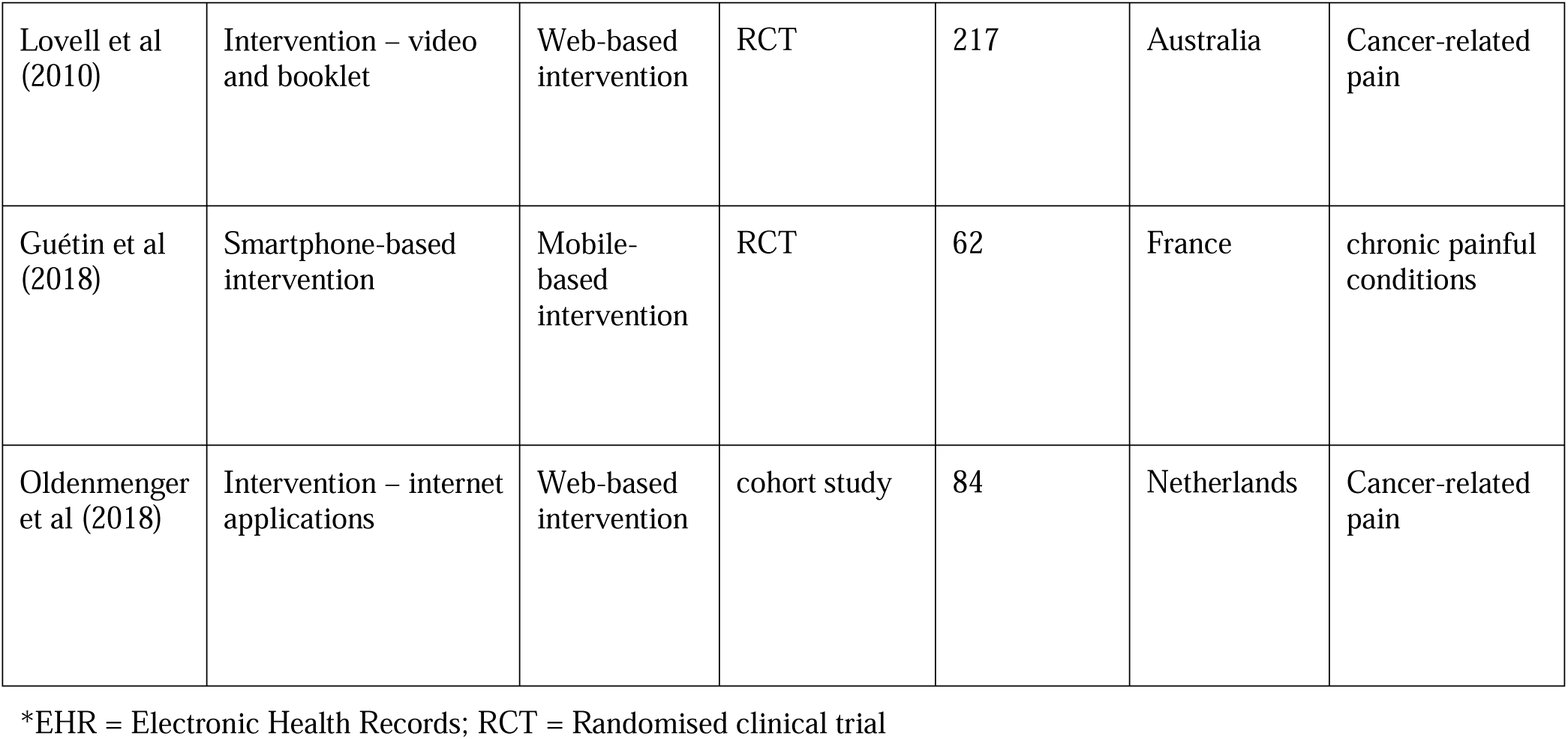
Characteristics of the systematically included studies

**Table 2.**
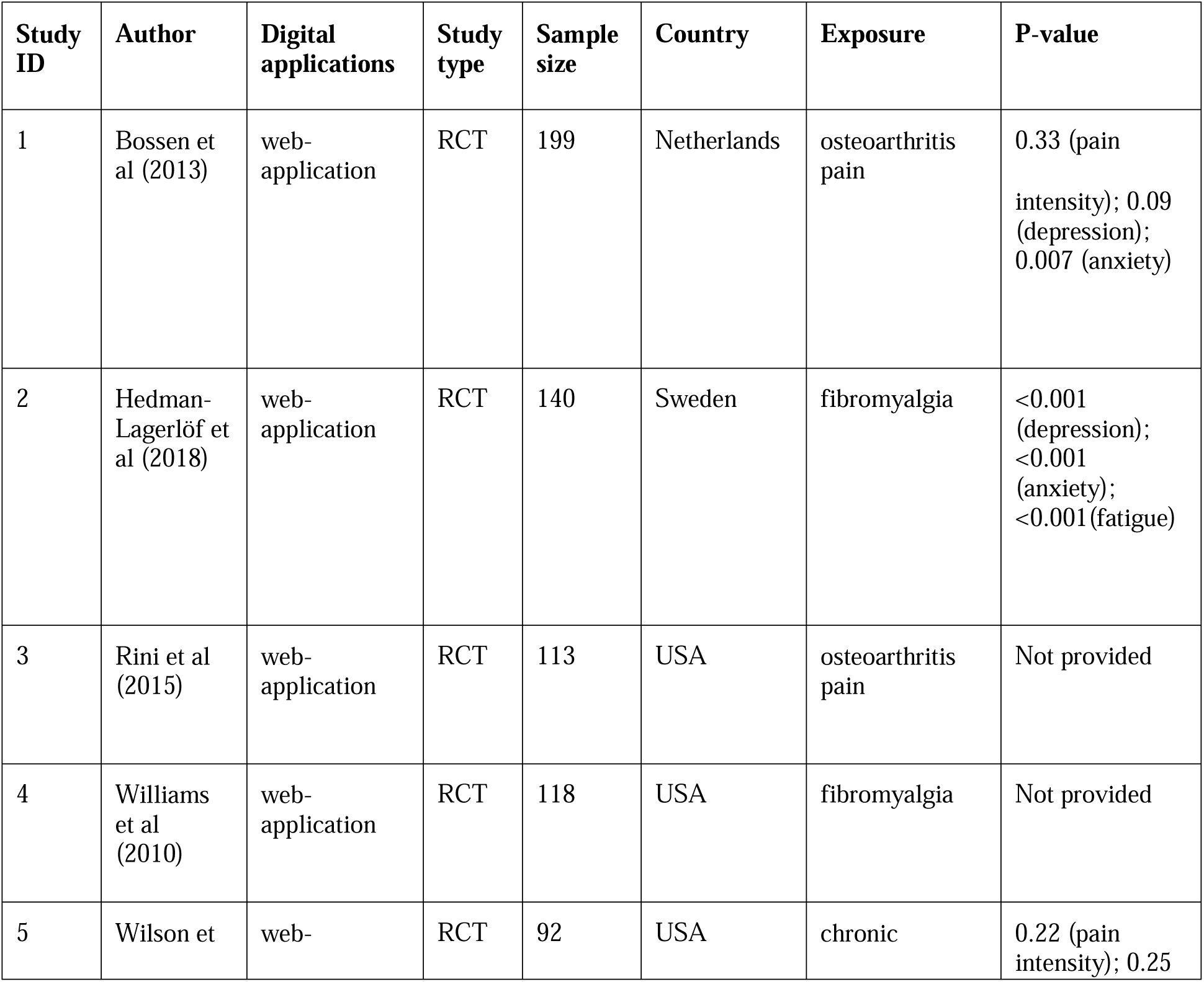
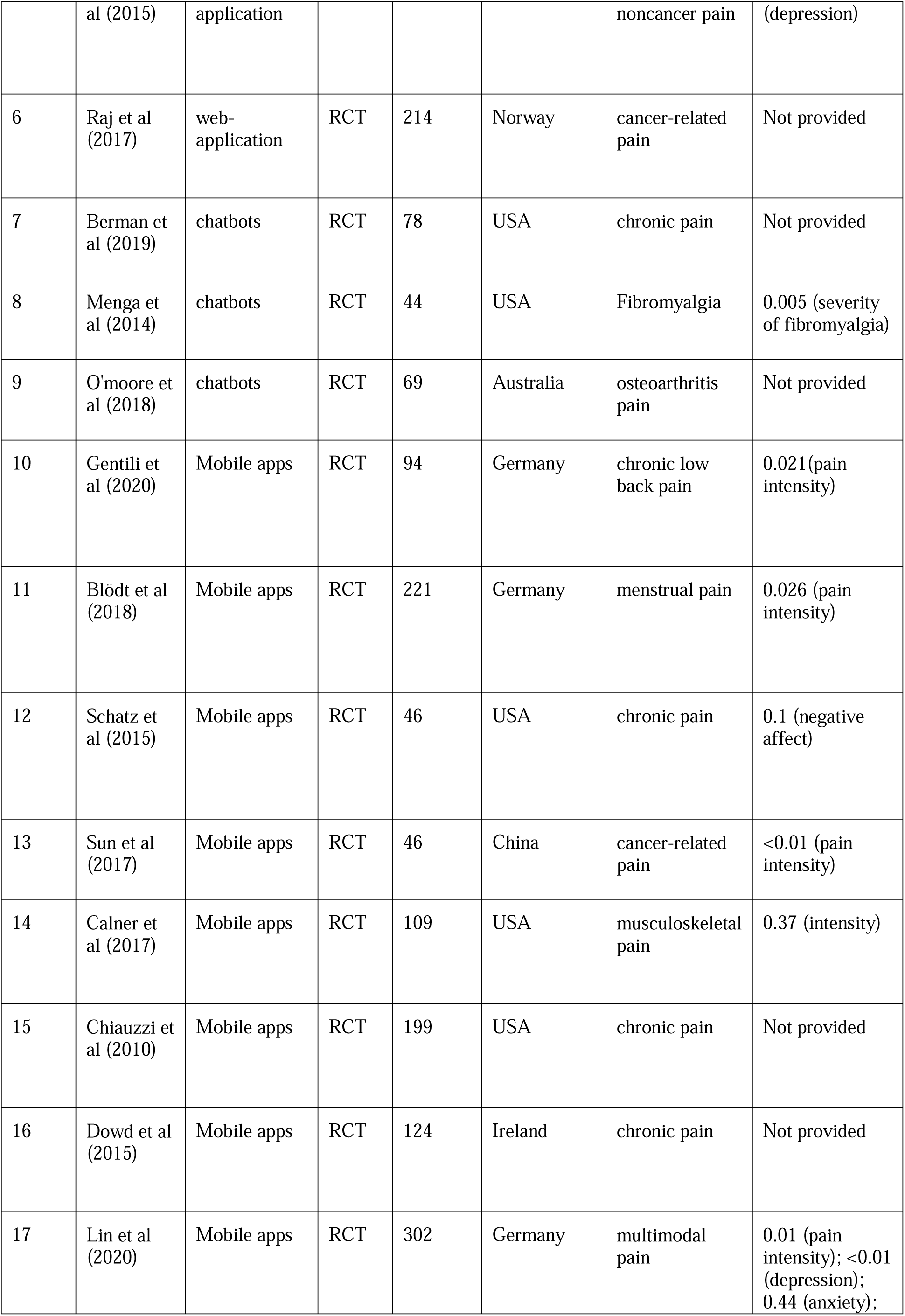
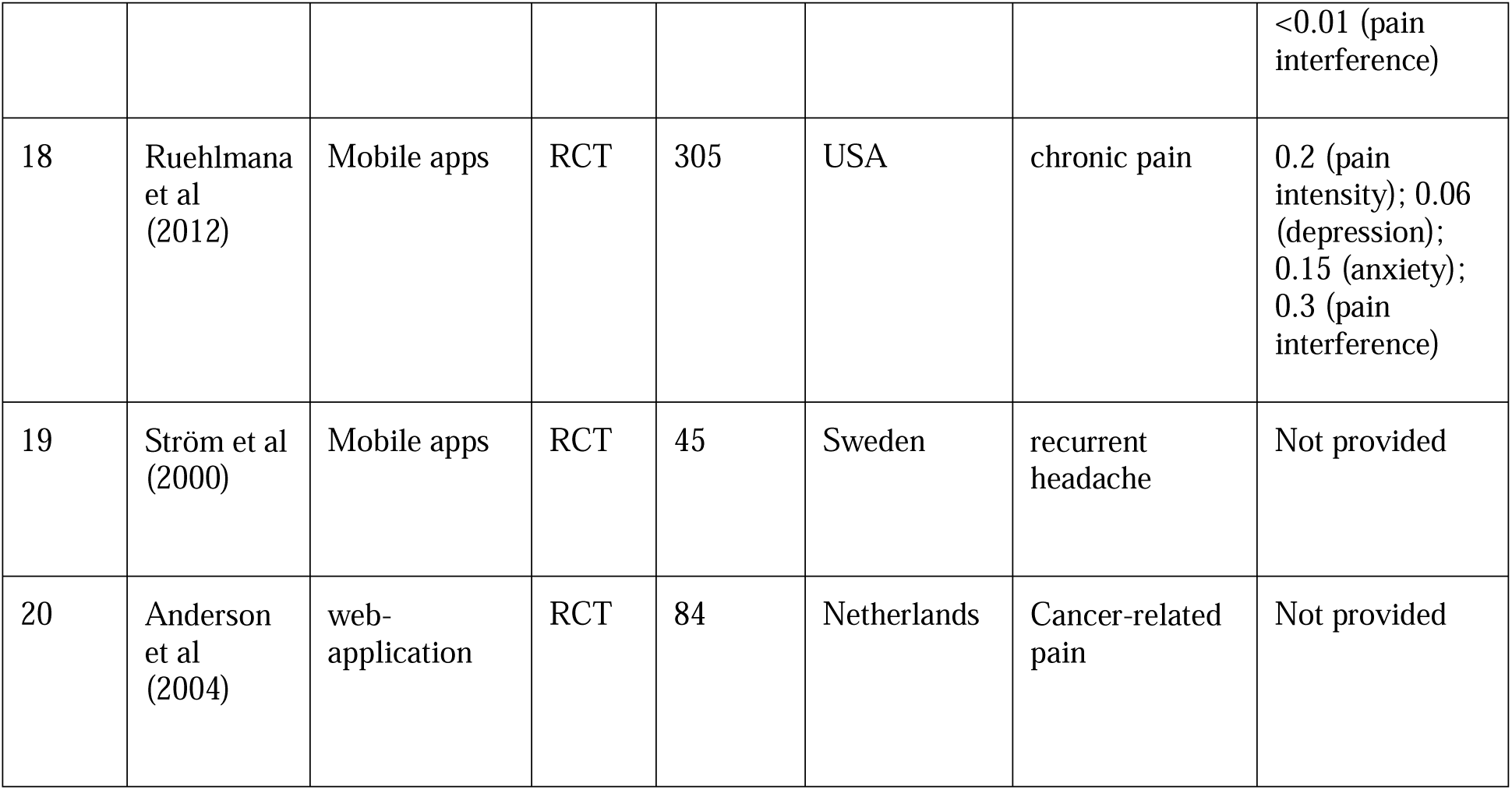
Studies included within the meta-analysis

**Table 3.**
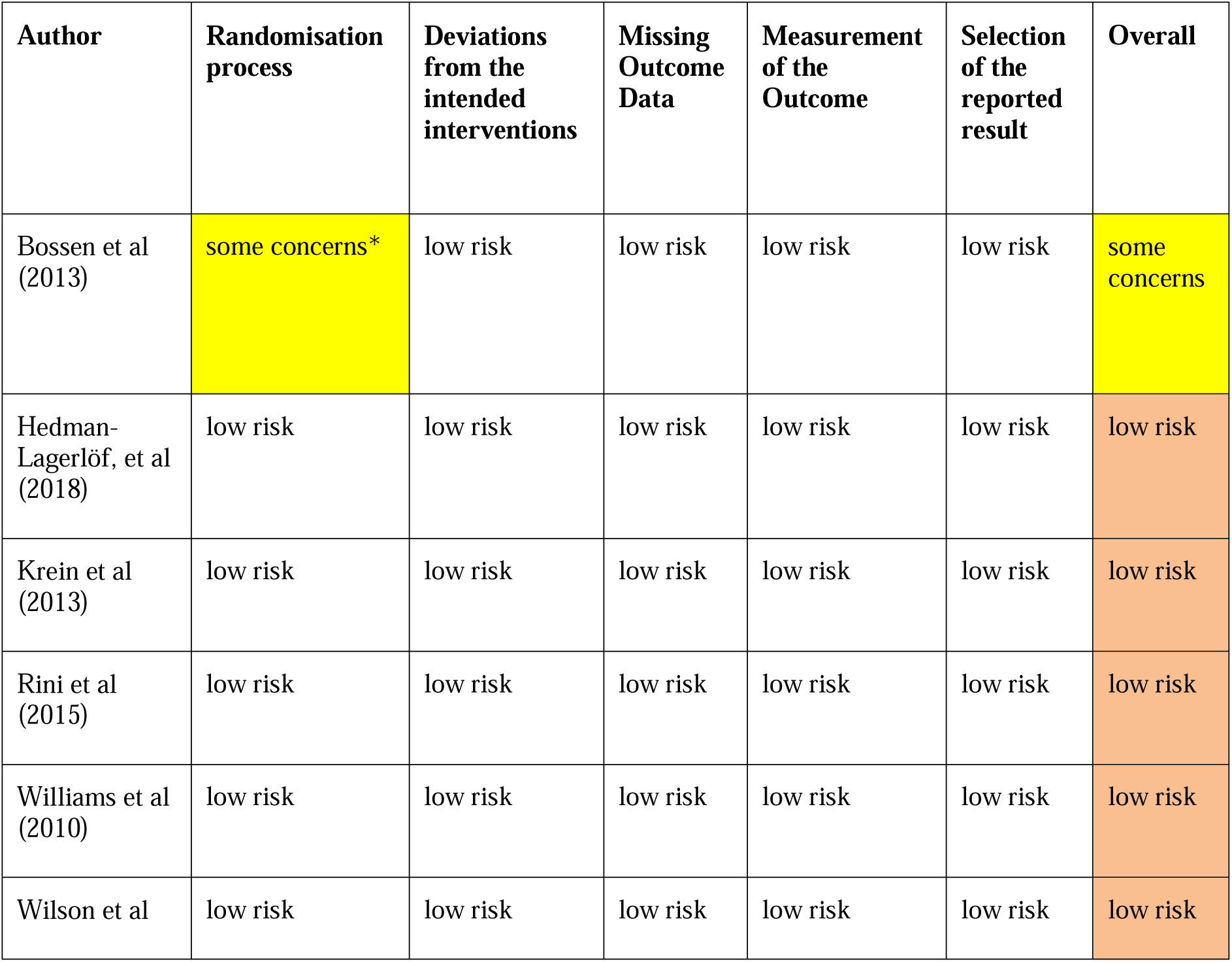
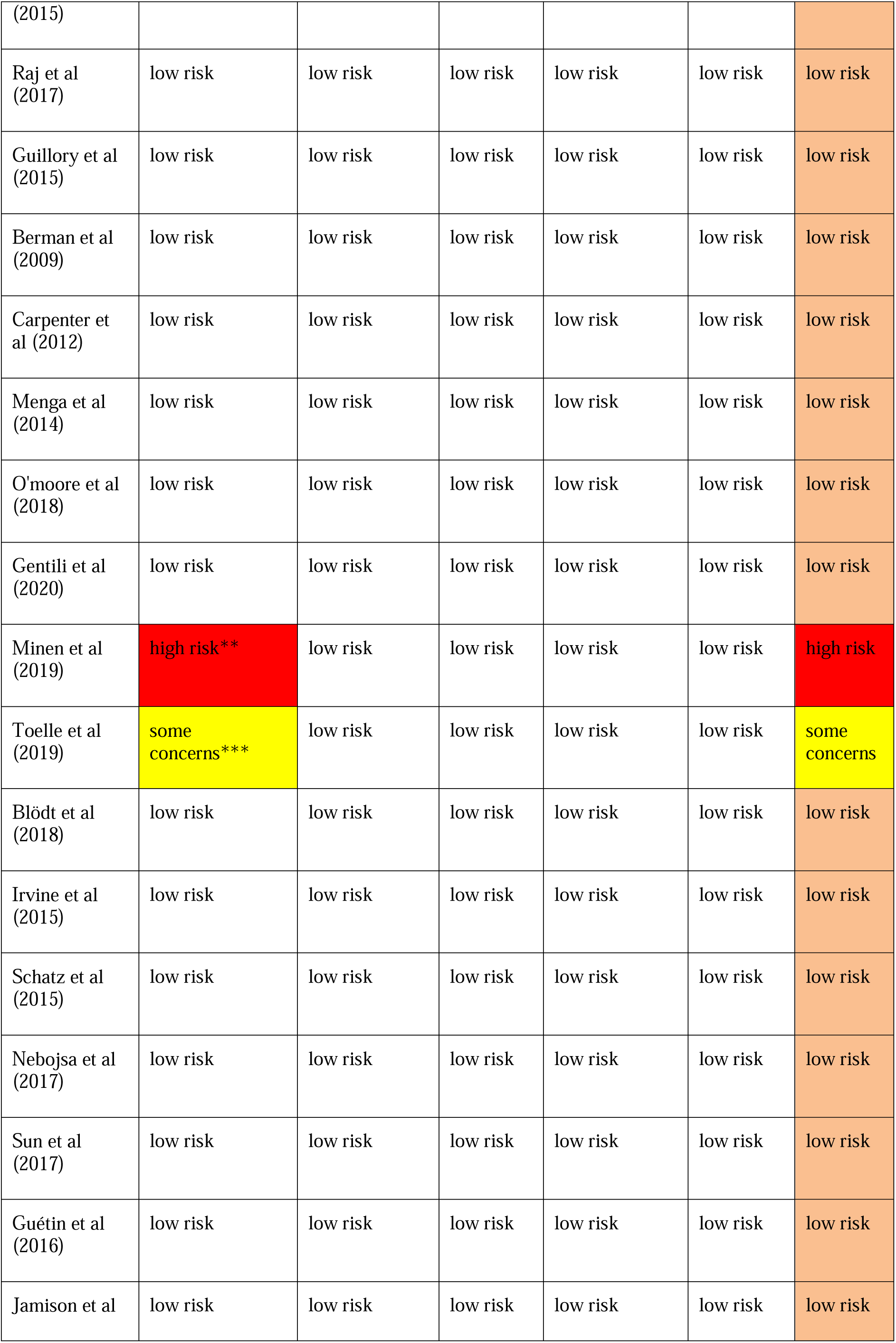
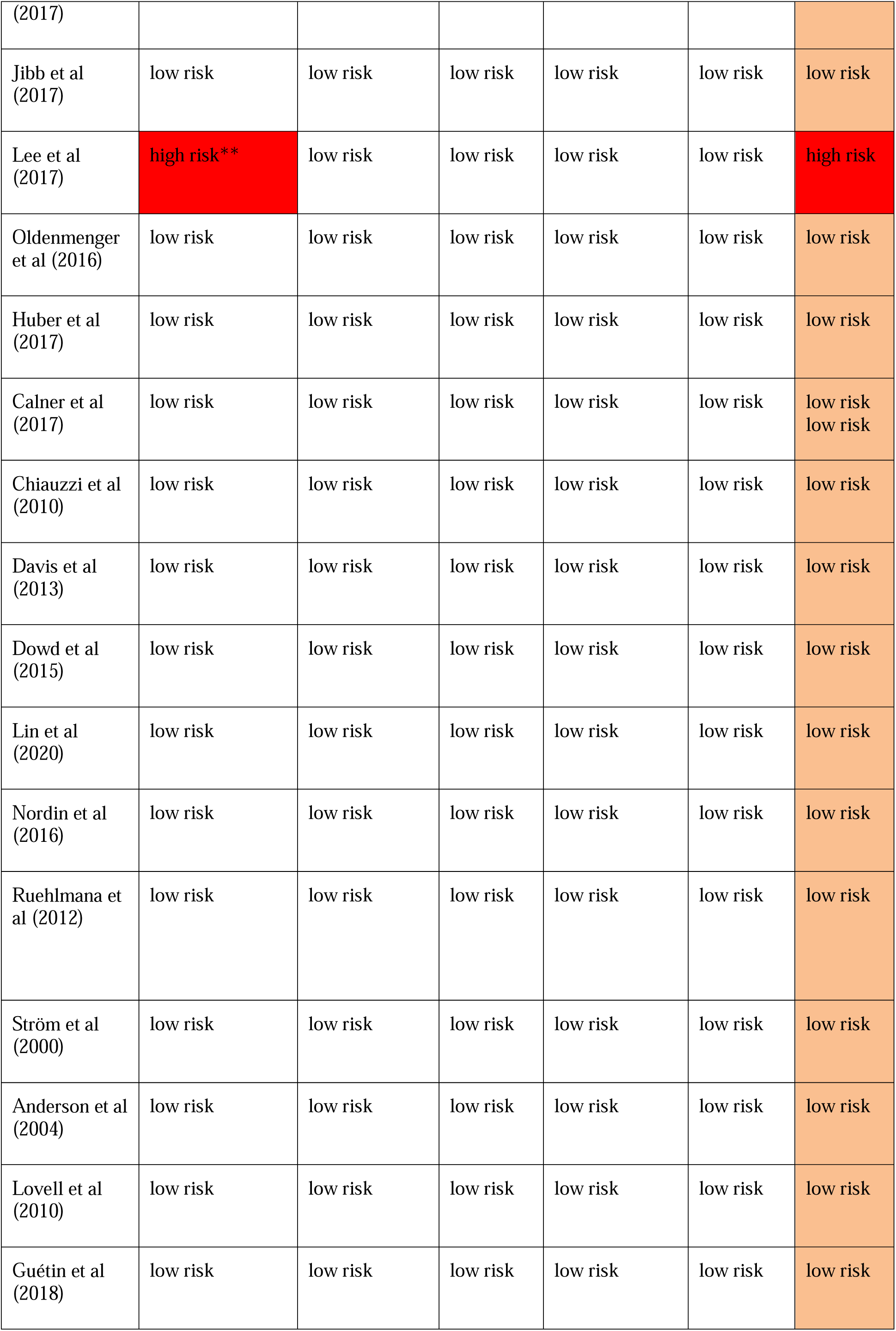
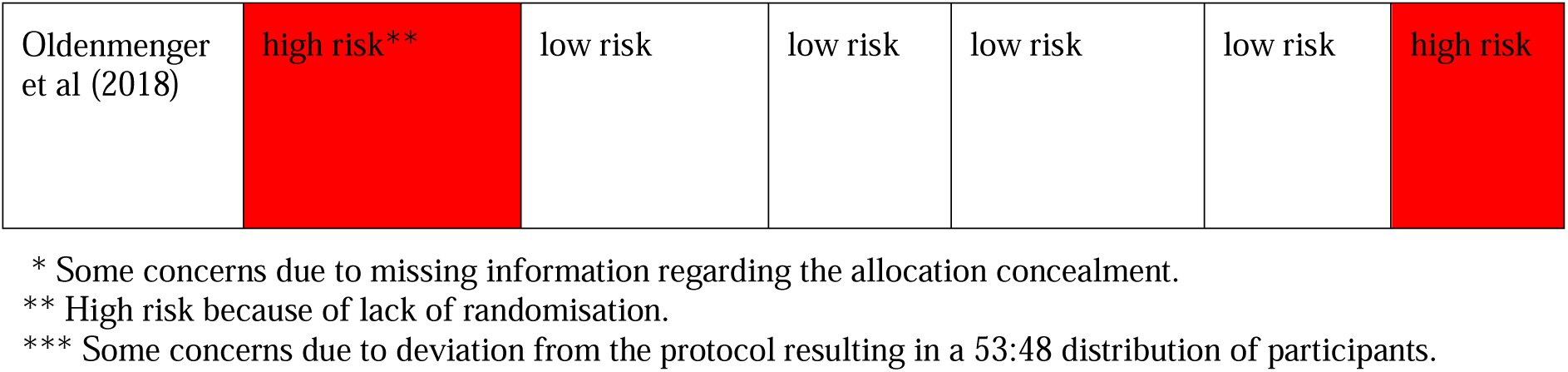
Risk of bias, according to the revised risk-of-bias tool for randomised trials (RoB 2.0)

### Meta-analysis

All 22 studies included in the meta-analysis reported more than one pain-related symptom. One primary outcome reported in 15 studies was pain intensity. 11 reported depressive symptoms and 9 anxiety symptoms. Pain interference was reported by 4 studies. Fatigue and sleep problems were included as secondary outcomes in two and one study respectively. Meta-analyses were conducted for each outcome separately.

### Pain intensity

All 15 studies provided the mean and SD. Therefore, the meta-analysis was based on the mean and SD. Figure 6 demonstrates a pooled SMD of 0.25 with a 95%CI of 0.03-0.46. SMD is statistically higher than 0; therefore, pain scores within the treatment group reduced compared to the control group, suggesting DM applications can significantly reduce symptoms of pain. A high heterogeneity of I^2^ =82.86% was identified for this group (*p*=0.02). A subgroup analysis was conducted to analyse the possible source of heterogeneity.

**Figure 6.**
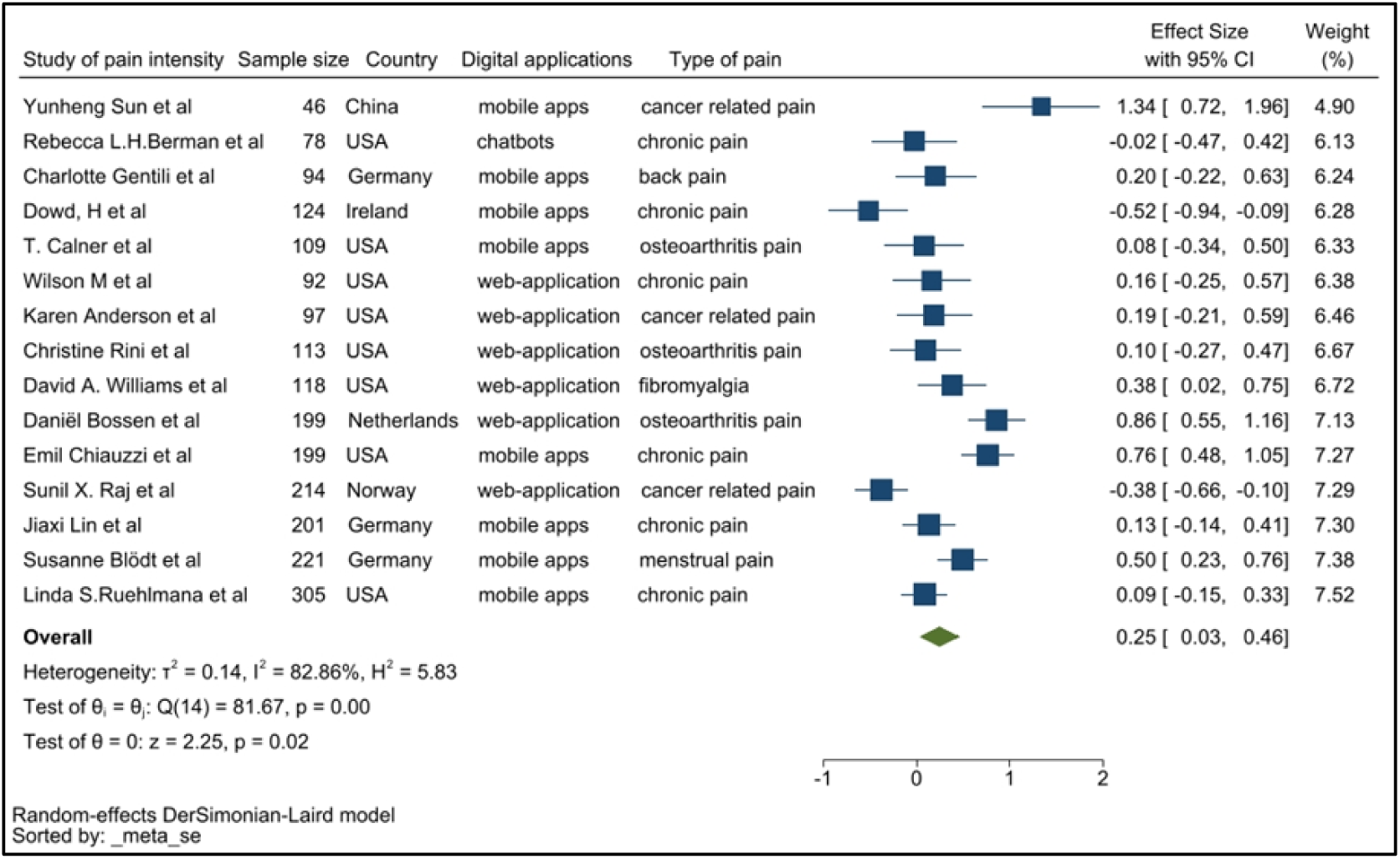
Forest plot of pain intensity

### Depression

The 11 studies reporting depressive symptoms used various assessment tools, including the Centre for Epidemiological Studies-Depression (CES-D), Beck Depression Inventory (BDI), Patient Health Questionnaire-8 and −9 (PHQ-8, PHQ-9), Hospital Anxiety and Depression Scale (HADS) and the Depression Anxiety Stress Scales (DASS). Ruehlman and colleagues (2012) used CES-D and DASS to assess the depression of the participants twice. To avoid duplication we used only one (CES-D) of the means and SD of these two assessments so that 11 studies were included in meta-analysis. Figure 7 showed that the pooled SMD was 0.30 with a 95%CI of 0.17-0.43, suggesting the use of DM applications reduced depression symptoms compared with the usual standard care, with an elevated heterogeneity of I^2^ = 34.72% (*p*=0.00).

**Figure 7.**
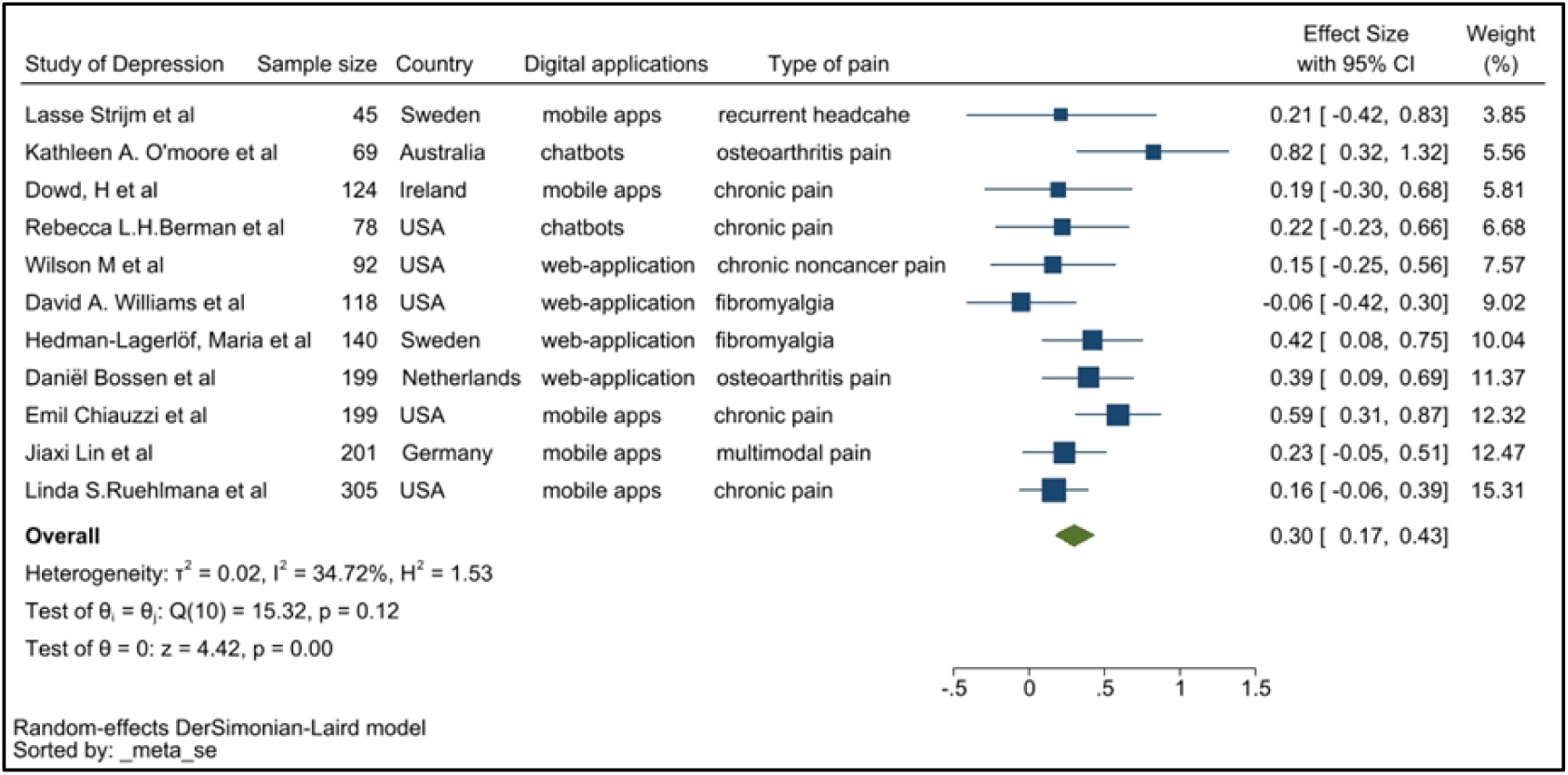
Forrest plot of depression

### Anxiety

Within the 9 studies reporting anxiety as a clinical outcome among chronic pain participants, the pooled SMD was 0.37 with a 95%CI of 0.05-0.69 (Figure 8). The SMD is significantly greater than 0, indicating anxiety symptoms among participants following use of DM applications improved more than the control group. Additionally, a treatment effect greater than 0 was seen in each individual study, thus each study concluded that DM applications improve anxiety symptoms compared with controls. Heterogeneity seen within this dataset was high with I^2^ = 88.34% (*p*=0.02), indicating a significant effect of the DM applications compared with the control group.

**Figure 8.**
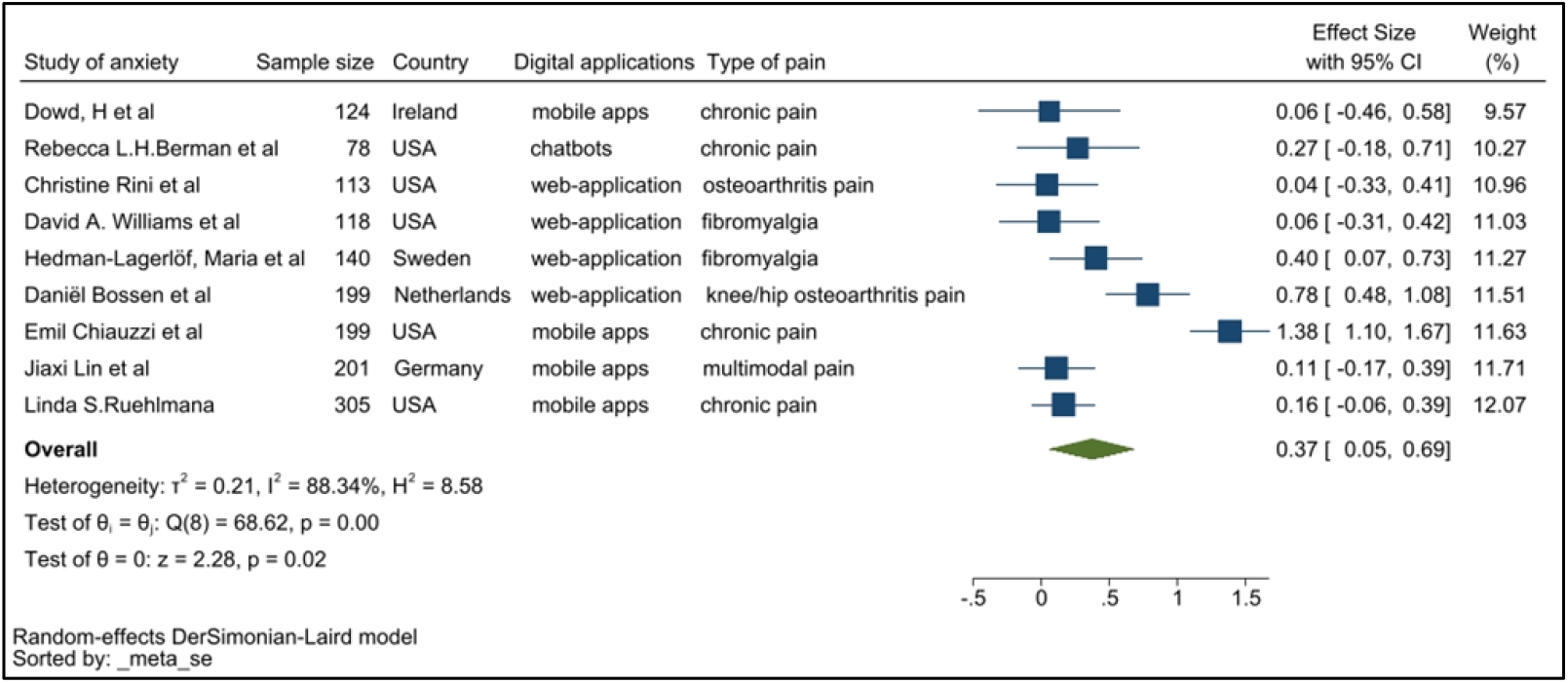
Forest plot for anxiety

### Pain interference

Four studies reported pain interference, an important outcome in pain research. Figure 6a demonstrates a pooled SMD of 0.15 with a 95%CI of −0.05–0.34. SMD was not significantly higher than 0, suggesting that the improvement within the treatment group was not significantly greater than control group. DM applications appear to have no effect on participants exposed to the application based on an, indicating mild heterogeneity. Therefore, a lack of a statistically obvious effect has been observed within the pooled dataset.

### Fatigue/sleep

Two studies reported on fatigue and one study on sleep issues. The forest plots for these factors are illustrated below (Figures 6b and 6c).

The pooled SMD for fatigue was 0.29, indicating the treatment group improved following the completion of the DM application use. However, this conclusion is not statistically significant given the 95%CI of −0.18-0.76. This could be due to the presence of only 2 studies, and more is needed to reach a conclusion.

The pooled SMD for sleep issues was −0.04 with a 95%CI of −0.4-0.32. This indicates that DM applications did not improve sleep-related issues and is of a lower score compared to the control group. However, to provide a more comprehensive conclusion to this phenomenon, further studies would be required.

### Subgroup analysis

Subgroup analysis was conducted to identify the source of raised heterogeneity when considering studies reporting on pain intensity. Initial analysis considered the categories of DM applications which included web-applications, mobile apps and chatbots. The analysis is demonstrated in Figure 7.

The pooled SMD for web-applications was 0.22, indicating web-applications could reduce the intensity of pain compared to the control group. The pooled SMD of mobile apps was 0.30. This demonstrates a larger effect size in relieving the intensity of pain compared to control groups and to those using web-applications. The pooled SMD for chatbots was −0.02, indicating chatbots have a limited effect in reducing the intensity of pain in patients compared to the controls. Heterogeneity remained high in all three subgroups, so a second subgroup analysis was conducted based on exposure of pain symptoms. The pain exposure sub-group analysis included identified specific pain conditions: fibromyalgia, back pain, chronic pain, osteoarthritis pain, menstrual pain, and cancer-related pain (Figure 8).

Heterogeneity could only be calculated in three of the subgroups. It remained high within these pooled subgroups, although at a lower level compared to previous analyses. Cancer-related pain reported the highest level of heterogeneity (I^2^=92.23%). Chronic pain and osteoarthritis pain groups reported an I^2^ of 82.44% and 85.19% respectively. However, due to the limited number of studies, pain and digital application exposures, the effect size could not be comprehensively assessed. A third subgroup analysis was conducted based on geographical locations (Figure 9a and 9b).

**Figure 9.**
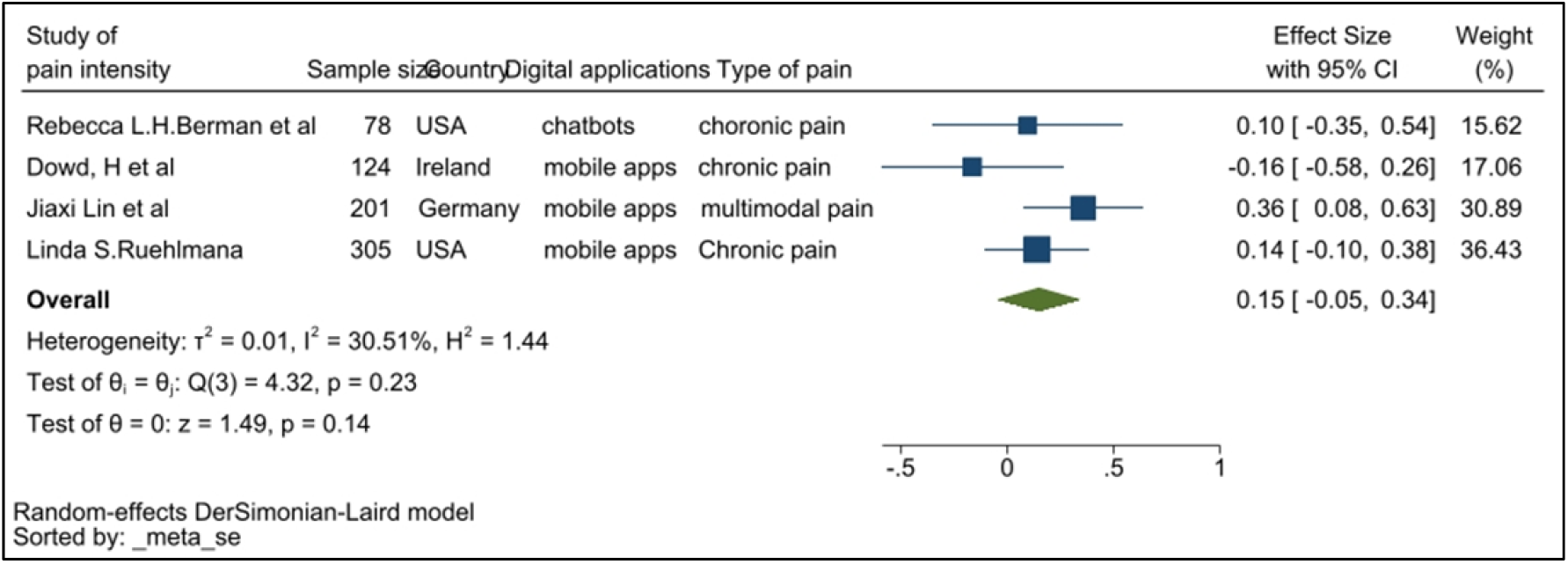
Forrest plot for pain interference

A sub-group analysis by country found that DM applications appear to be effective within populations in America, China, Germany, and Netherlands, while for Ireland and Norway, a statistically significant effect was lacking. Only mild heterogeneity levels were indicated for America (I^2^=61.54%) and Germany (I^2^=45.91%). The heterogeneity may well be due to nationality and ethnicity.

### Sensitivity analysis

Based on the meta-analysis and the sub-group analysis conducted to demonstrate pain intensity outcomes from the digital tools reported by Anderson et al,^49^ Chiauzzi et al^42^ and Sun et al,^34^ the standard mean deviation (SMD) was high. The primary populations of Chiauzzi and colleagues (2010) and Anderson and colleagues (2004) were African American followed by Hispanic, whilst Sun et al (2017) reported on a population of Chinese patients. Similar ethnicity and race patterns were found among 12 of the 15 studies in the meta-analysis. Of these, 5 reported ethnicity, although over 50% of the sample size was Caucasian. The other 7 did not provide specific percentages of the Caucasian representation. A sensitivity analysis was conducted to assess ethnic variability within the pooled sample size, which resulted in a SMD of 0.14 with a 95%CI of −0.07-0.35 (Figure 10).

**Figure 10.**
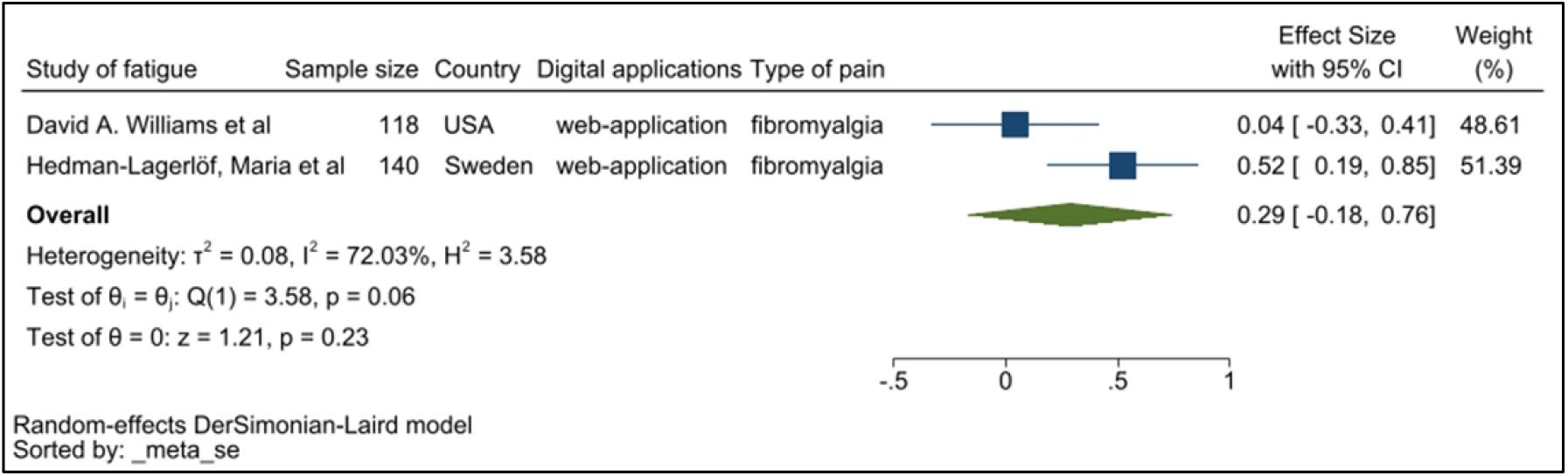
Forest plot of fatigue

The sensitivity analysis reveals DM applications appear to benefit patients. However, to conclusively demonstrate a statistical significance more studies would be required. The *p*-value where the reported SMD was greater than 0 was 0.2, indicating there is a 90% probability that the DM application would have a positive impact on the patient’s pain. It is equally vital to recognize that the predominantly African American and Hispanic population-based studies reported a SMD of 0.76 with a 95% of 0.48-1.05, and a study consisting entirely of African American and Hispanic participants reported a SMD of 0.19 (95%CI −0.21-0.59). Sun et al (2017) reported a SMD of 1.34 with a 95% CI of 0.72-1.96. Therefore, DM applications appear to have a positive impact on patients.

The sensitivity analysis shown in Figure 16 demonstrates strong heterogeneity. The main source of heterogeneity could be the difference in the treatment interventions deployed by way of the DM application. As this is associated with the interventions themselves rather than the DM applications, it is beyond the scope of this study, and could be explored in the future. Pooled SMD of web-application, mobile apps and chatbots were 0.22, 0.1 and −0.02 respectively. Figure 11 demonstrates that the most effective DM application could be mobile apps since web-applications are not a self-reported method.

**Figure 11.**
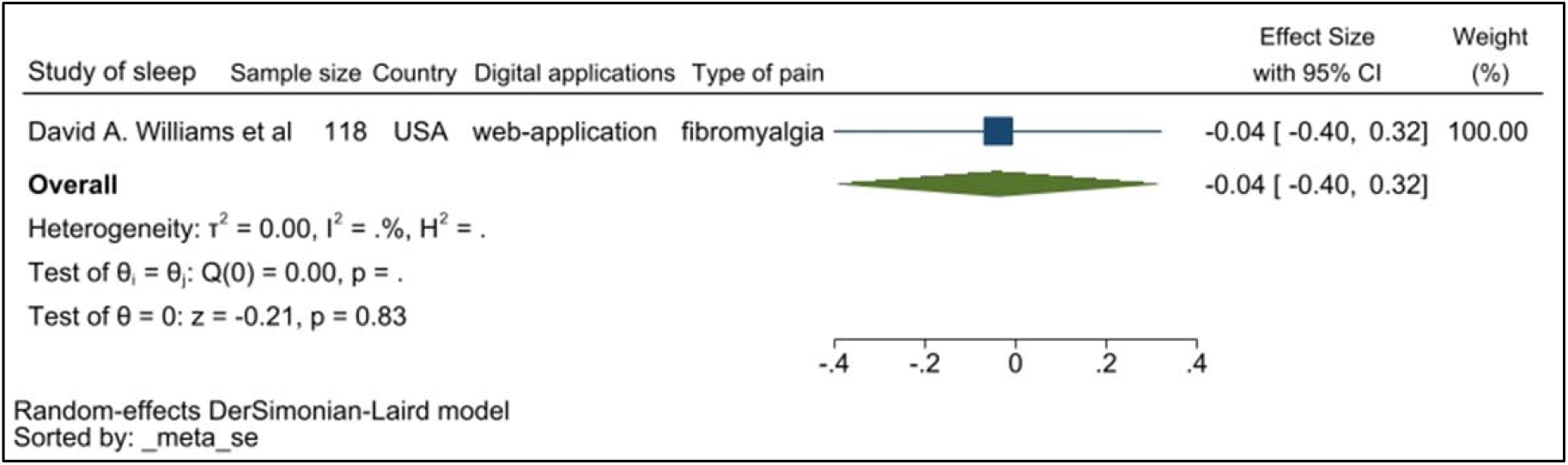
Forrest plot of sleep

### Publication bias

Publication bias was assessed and reported using funnel plots and Egger’s test to examine the small-study effect. Publication bias appears to be smaller among studies associated with fatigue and sleep, and higher in studies demonstrating pain intensity, depression, anxiety, and pain interferences. There is a lack of significant publication bias based on the funnel plot (Figure 12a, below). The Egger’s test *p*-value is 0.932, indicating the lack of a small study effect. However, there are 5 studies that fell outside the 95%CI which could affect our detection of publication bias. The *p*-value is not high but it is limited by the data and experimental quality.

**Figure 12.**
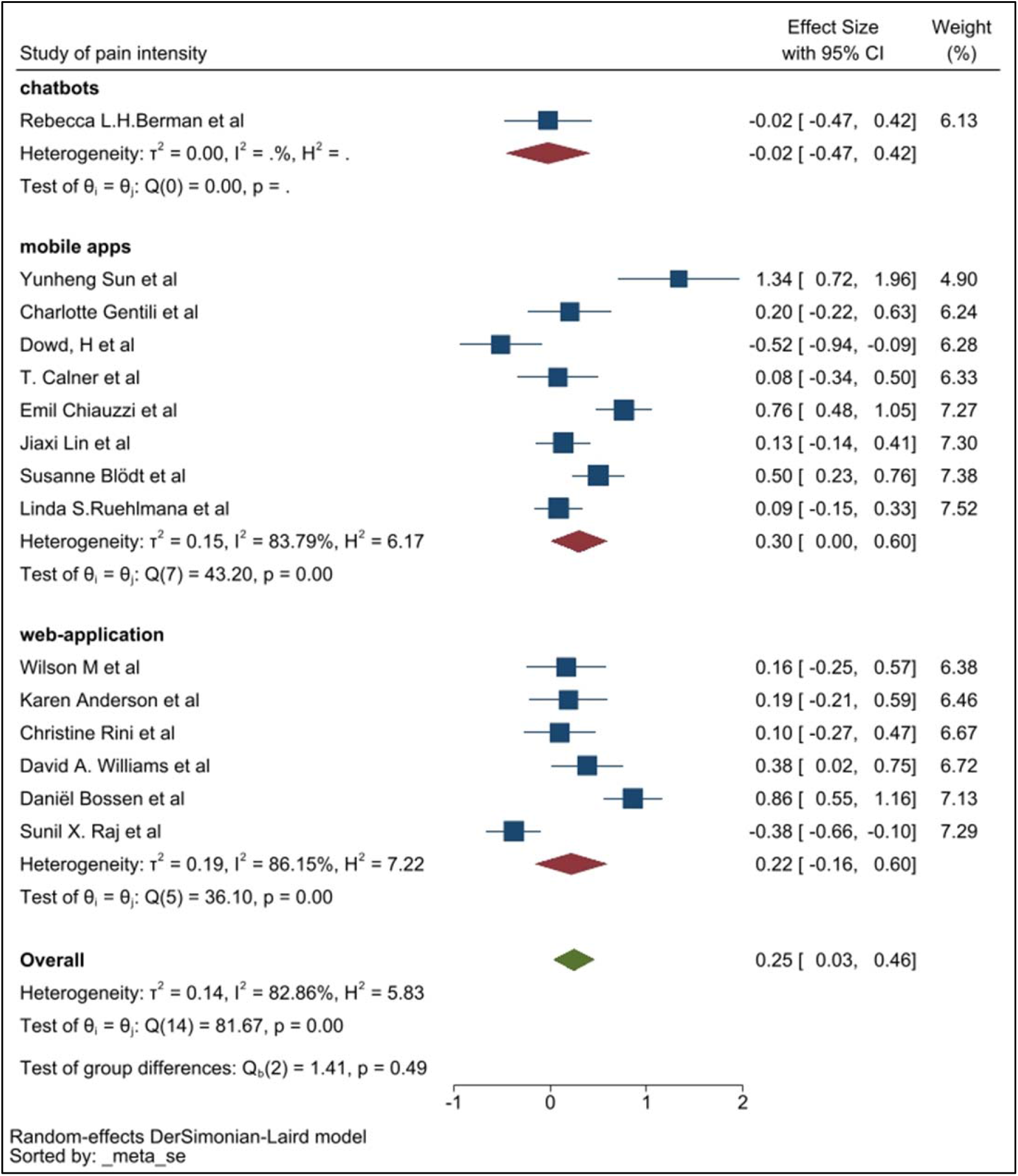
Subgroup analysis for pain intensity (web-application, mobile apps, chatbots)

**Figure 13.**
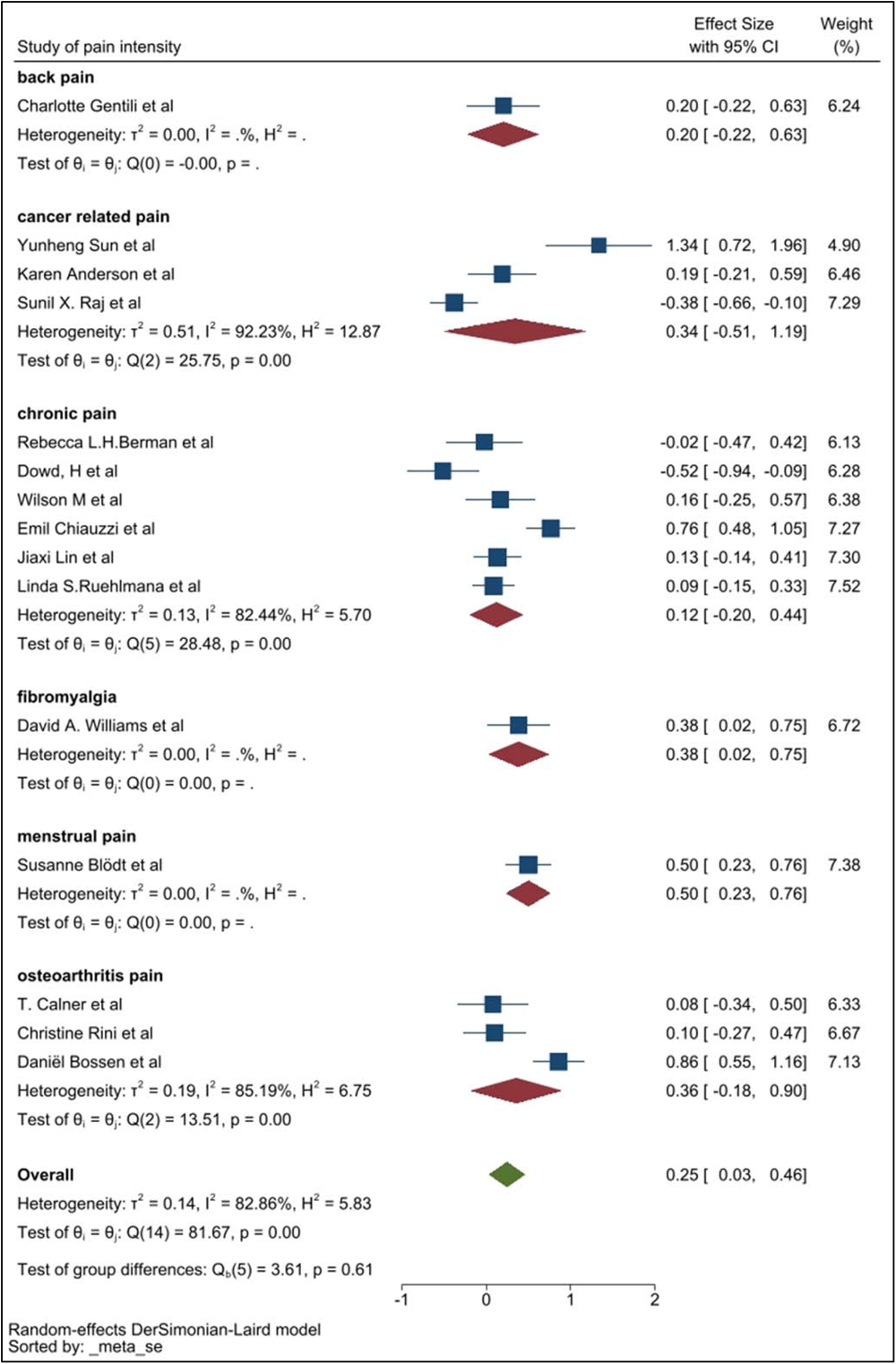
Subgroup analysis for pain intensity (by pain type)

**Figure 14a.**
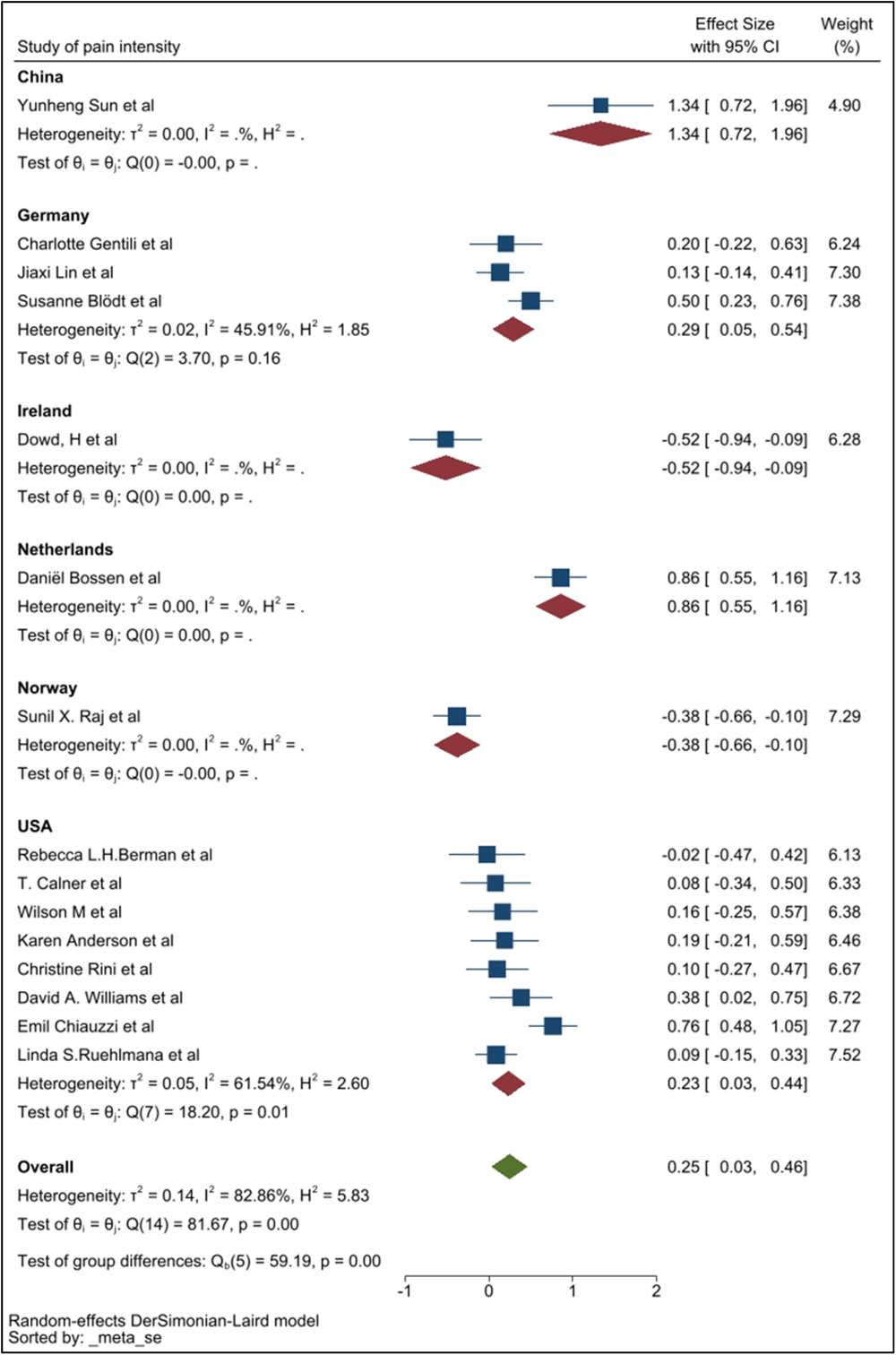
Subgroup analysis for pain intensity (by country)

**Figure 14b.**
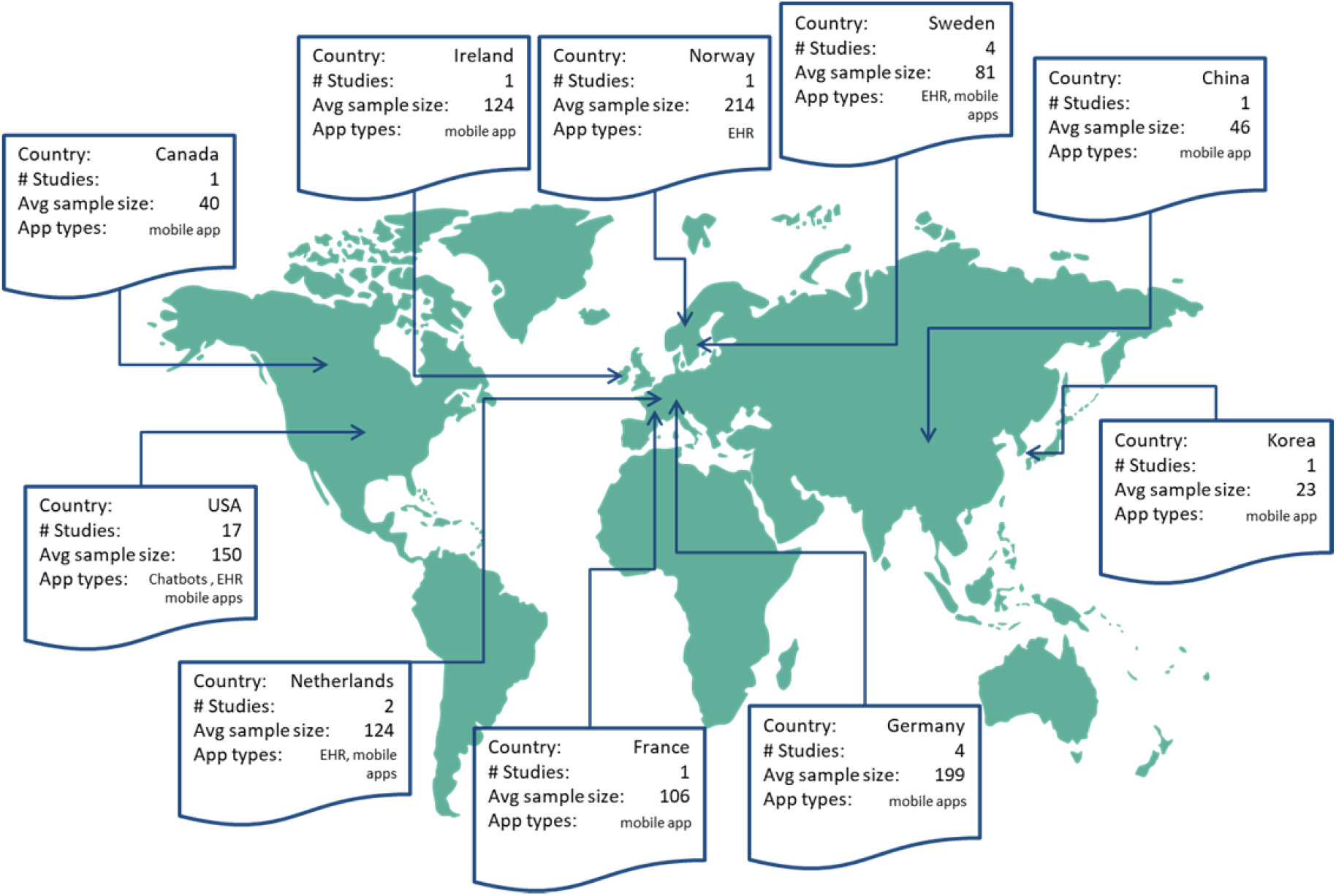
Geographical spread of data collected for the systematic review

**Figure 15.**
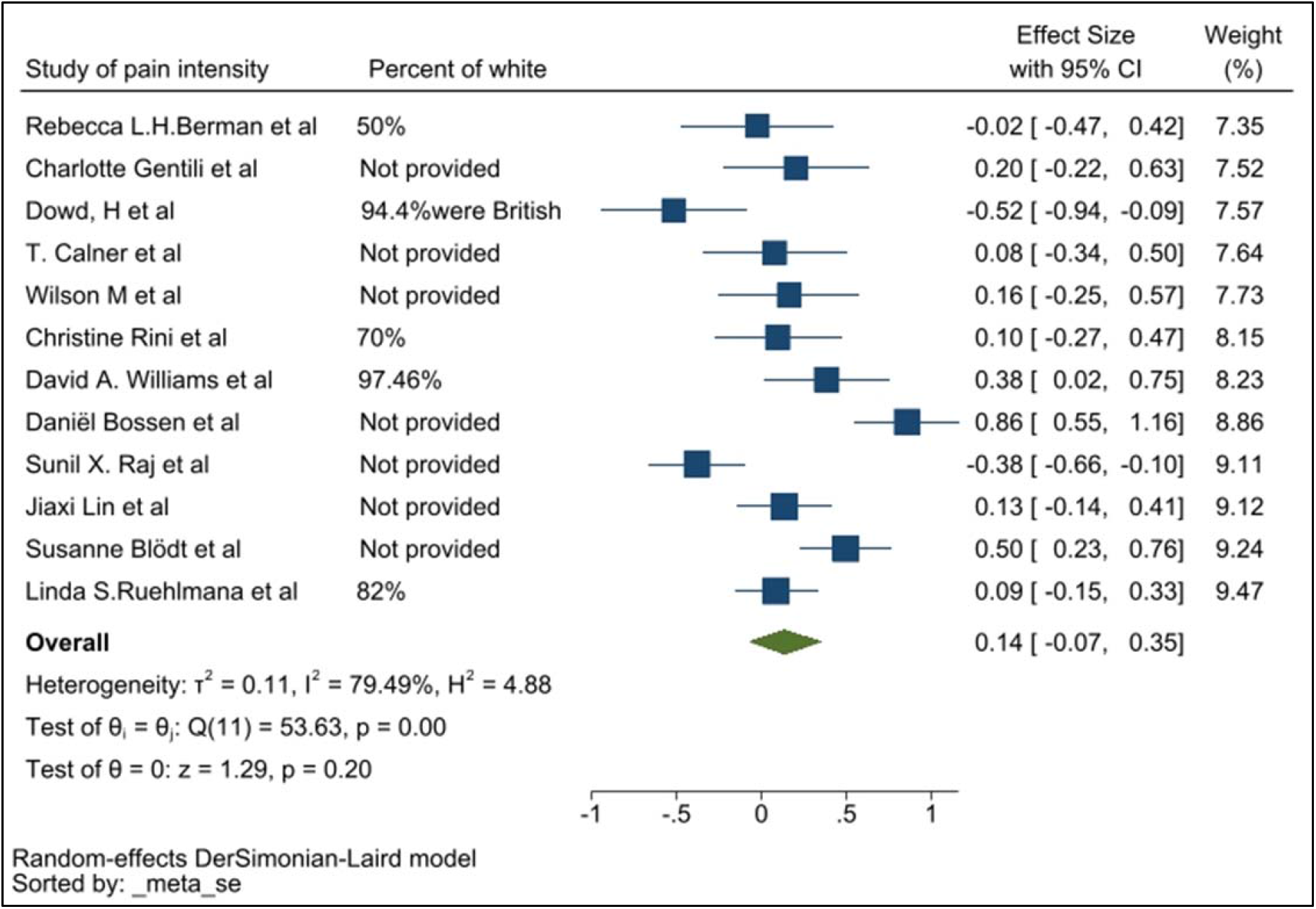
Sensitivity analysis without 3 BAME studies^33,41,60^

**Figure 16.**
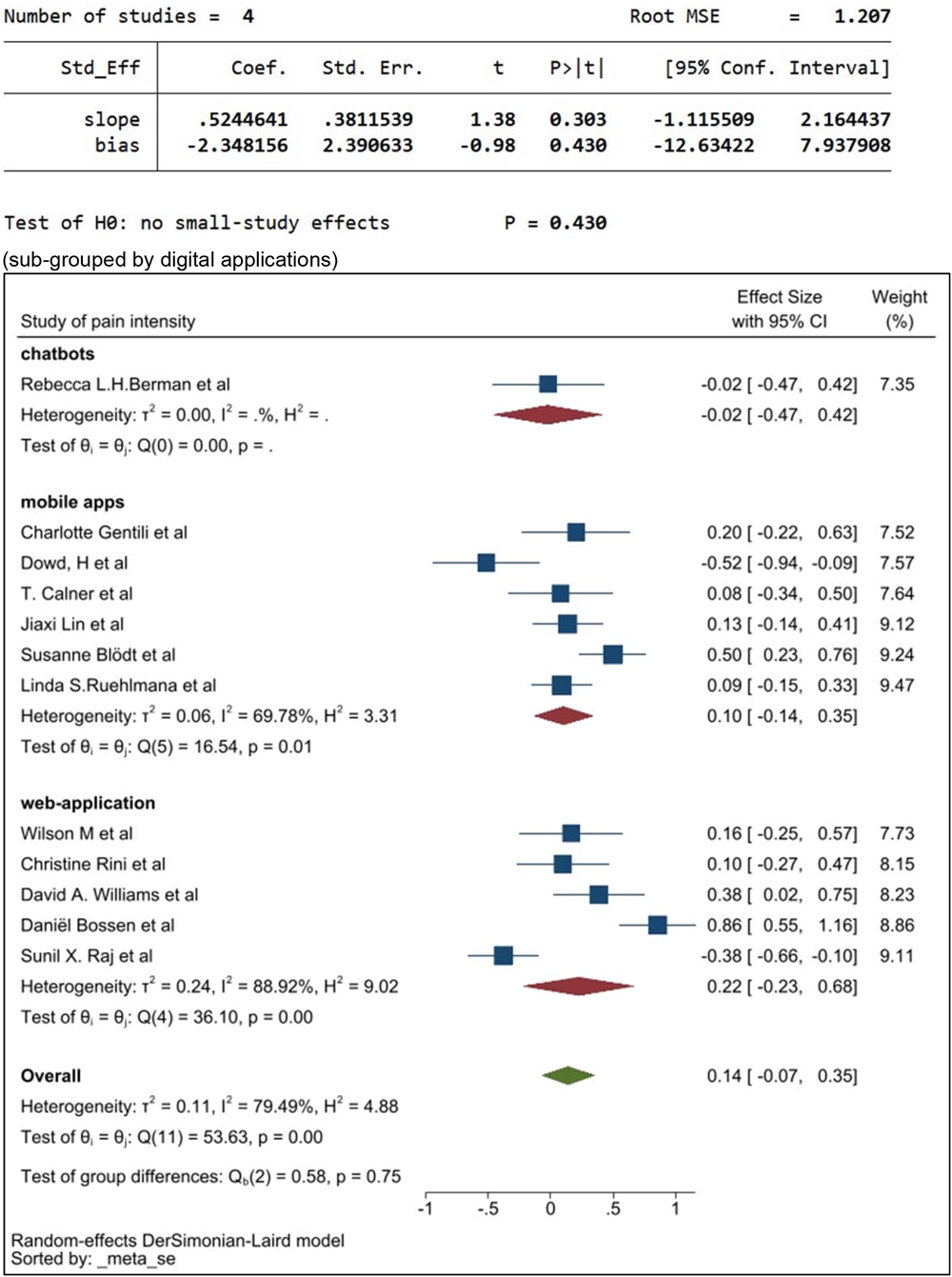
Sensitivity analysis without 3 BAME studies^33,41,60^

**Figure 17.**
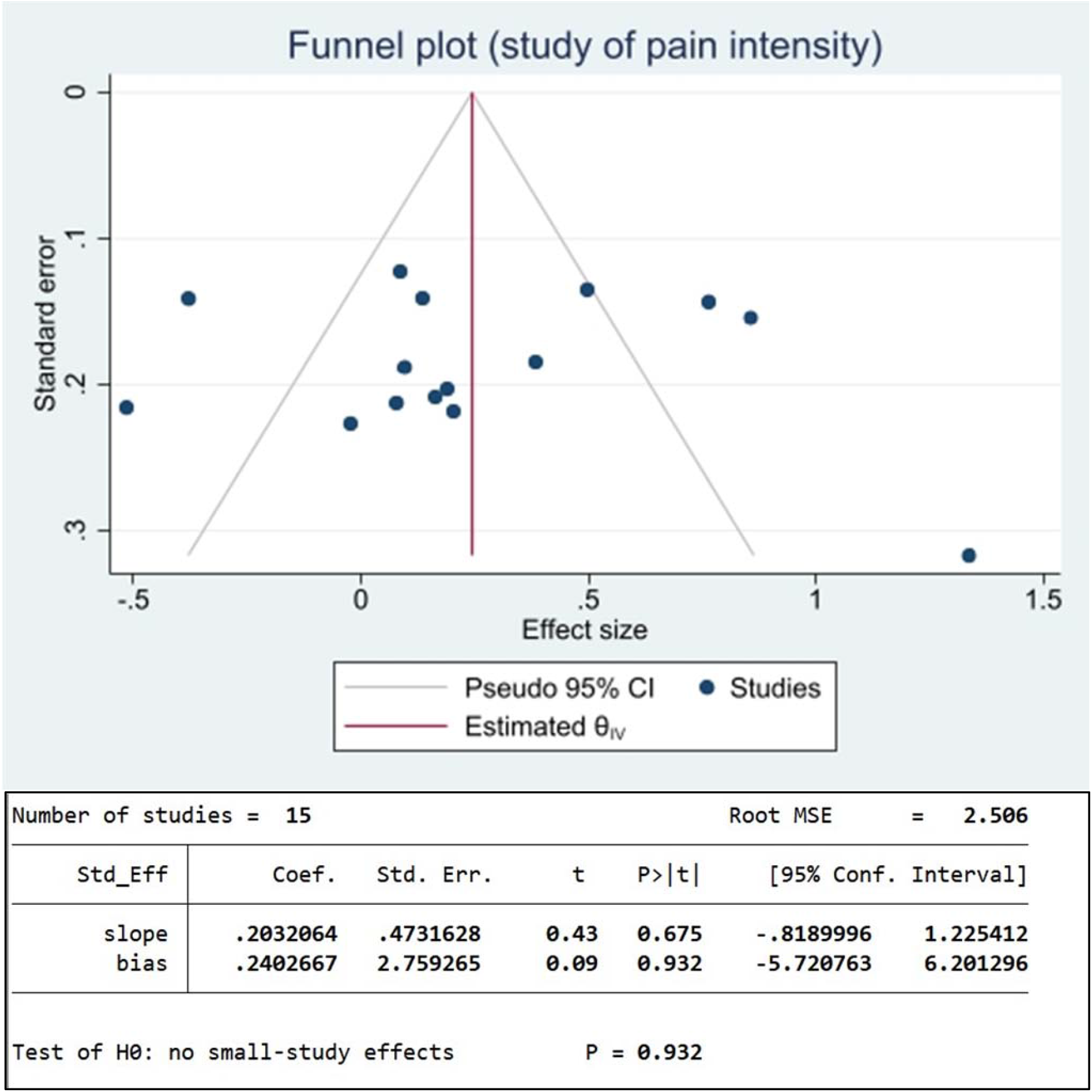
(a) funnel plot for pain intensity; (b) egger’s test for pain intensity.

**Figure 18.**
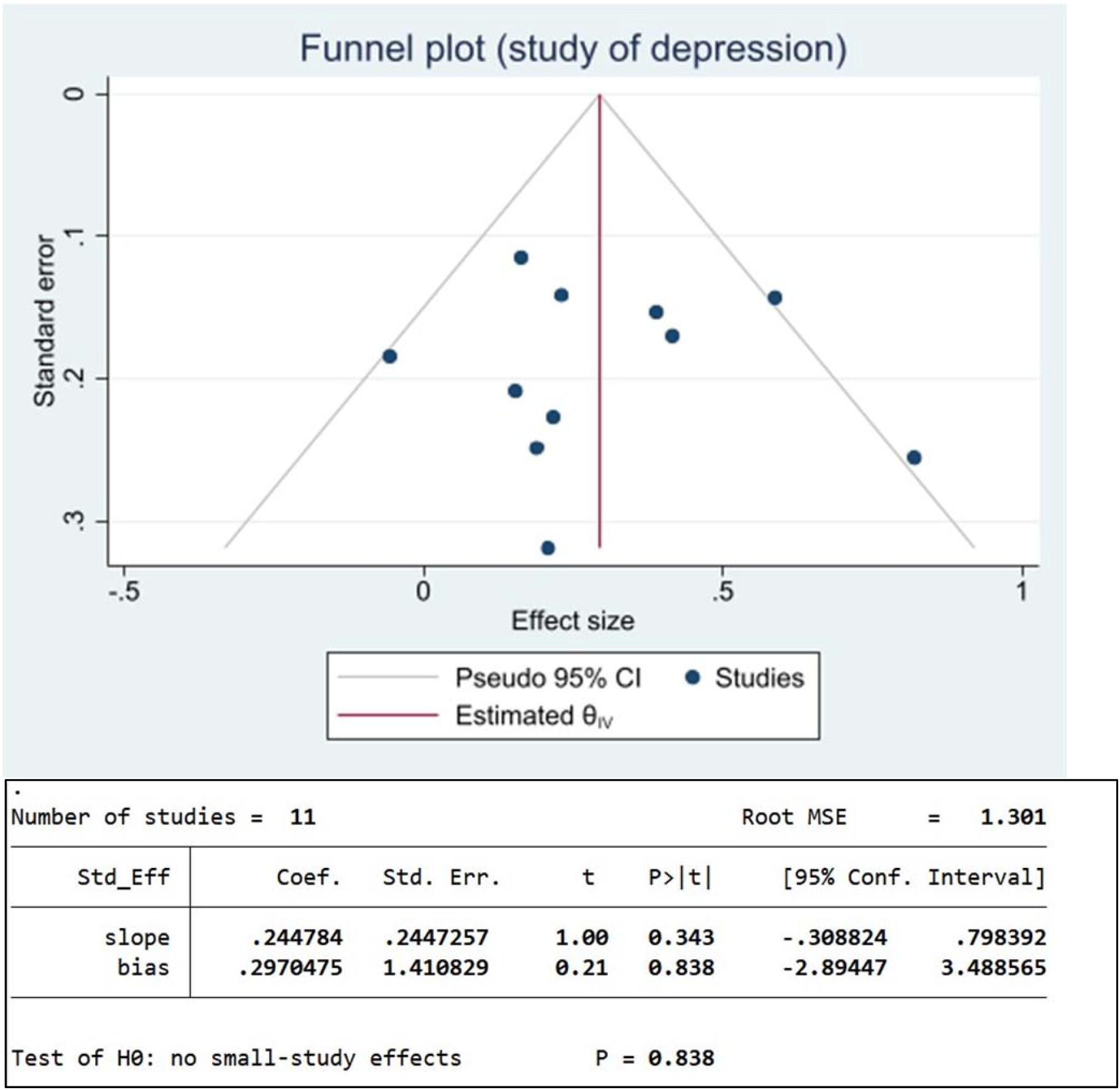
(a) funnel plot for depression; (b) egger’s test for depression.

**Figure 19.**
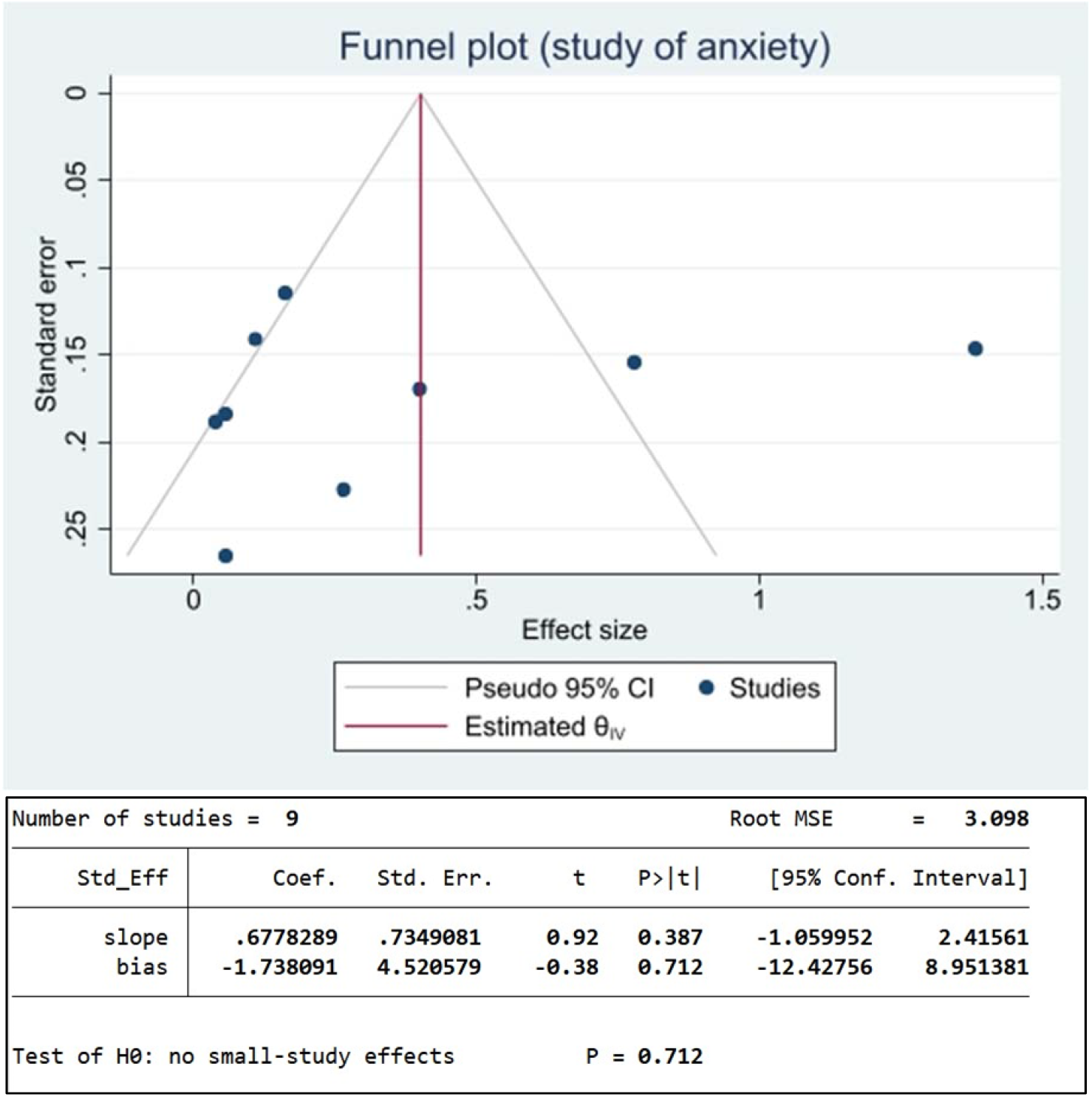
(a) funnel plot for anxiety; (b) egger’s test for anxiety.

**Figure 20.**
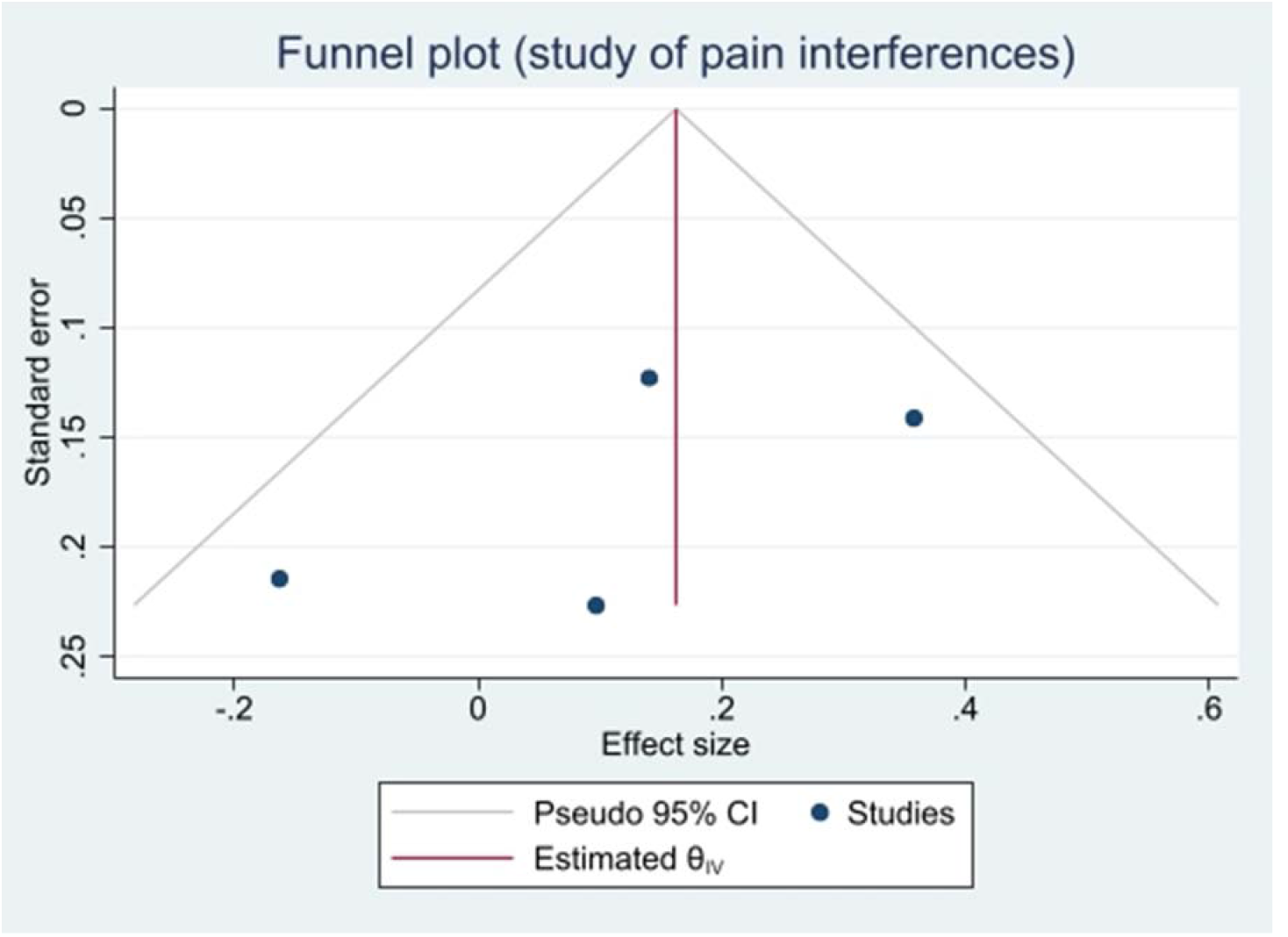
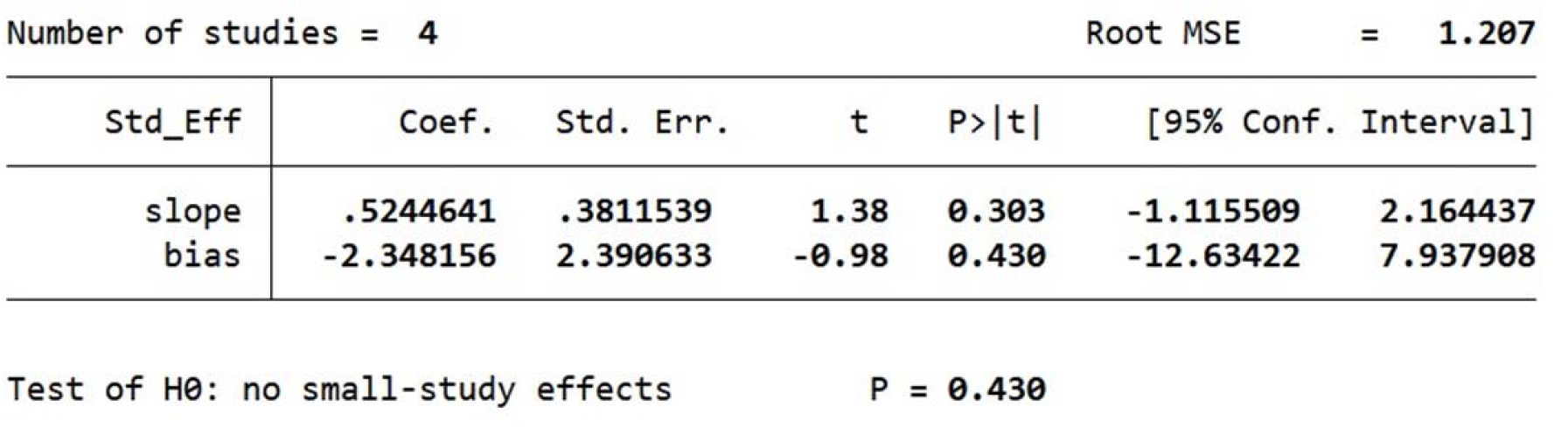
Funnel plot for pain inferences

Figures 12b and 12c indicate a lack of publication bias statistically for depression and anxiety, with Egger’s test *p*-values of 0.838 and 0.712 respectively. Pain inferences (Figure 12d), which was included in four studies, had an Egger’s test *p*-value of 0.43, which demonstrates we cannot detect a publication bias. The low numbers of studies reporting outcomes for fatigue and sleep problems meant analysis of publication bias was not possible.

## Discussion

The prevalence of DM applications within pain research appear to be moderate and is focused around developed countries such as America and Germany. China appears to be the only country within Asia to have conducted a study to assess the use of DM applications among patients with chronic pain. This demonstrates an urgent need to conduct evaluations of these DM applications in low-income and middle-income countries to optimise and evaluate the efficacy and acceptability among patients and clinicians. Patient-reported DM applications identified in the systematic review could be categorised primarily as mobile apps and chatbots, as EHR systems were used to assess pain-associated outcomes. As a result of these differences, the prevalence of DM applications was meta-analysed at a granular level to identify and report pain outcomes such as depression, anxiety, pain intensity and pain inference that were assessed by the tools. The lack of uniformity among the assessments used within the applications are another issue that requires further elaboration if these are to be used by clinicians as part of a patient’s ongoing clinical management. The assessments used also appear to be non-specific to a particular group of patients. Often, the studies did not report on underlying conditions or if the pain conditions had a clinical diagnosis. Thus, it is challenging to demonstrate that users demonstrated true clinical benefit. This suggests there is little quantifiable data to provide a comprehensive conclusion in terms of the generalisability and feasibility of these applications globally.

We identified multiple themes and sub-themes in this analysis that were pooled as mobile applications, EHR and chatbots. Mobile applications have grown rapidly to support the management of pain disorders such as migraine, back pain and fibromyalgia by offering educational components, exercise platforms, relaxation techniques and mindfulness-based options to name a few. These options provide feedback and allow engagement and adherence of the users. This may explain why mobile applications demonstrated better results compared with other DM applications in the management of chronic pain. Another facet to consider is the inclusion of these datasets to maintain a structured approach to deliver effective continuity of care provision. Trials promoting the evaluation of data in a comprehensive manner through systems that allow the standardisation and acceptance of quality data would increase the acceptance of digitised data. Trials involving DM applications that incorporate AI-based clinical algorithms to assist with the evaluation of pain and outcomes in patients with cancer appear encouraging.^53^

Ledel Solem and colleagues (2019) reported adult participants were in favour of using DM-based self-management interventions for chronic pain management.^54^ Patients felt that the accessibility, usability, and personalisation were vital for DM tools, and suggested that these should be further developed to distract them from pain, regardless of pain intensity and cognitive capacity.

The benefits of harnessing DM within the context of pain medicine could improve both clinical and patient-reported outcomes. Evidence-gathering to support therapeutic efficacy for pharmacological or surgical treatments requires effective and robust methodology, yet rigid traditional trial designs remain inefficient and struggle with implementation into clinical practice, limiting sustainability within healthcare systems. Computer-based technology could address these obstacles in research. The flexibility and accessibility of digital technology enables a more convenient and improved consenting process. This could allow easier enrolment and participation in studies for populations disadvantaged by mobility or literacy issues. Increased recruitment and retention lead to larger study populations with greater data validity, and aids researchers by speeding up recruitment and assessment of large trial populations.

Digital clinical trials are key in collating all the above factors, as it is a fundamental tool in assessing the efficacy and safety of novel drugs, medical devices, and health system interventions. Traditional clinical trials have demonstrated the validity, acceptability, and sustainability of the interventions, whilst digital clinical trials could leverage technologies to engage and report trial-specific measurements associated with the interventions being tested at a lower cost.^59^ Conceptualising digital clinical trials for pain medicine could have added benefits, especially for patients who could report pain episodes daily. That would allow digital analytics to assess considerations clinicians need to make when developing clinical treatments. Additionally, data science approaches could be leveraged in this instance to develop novel clinical methods to best utilise trial data with ‘real-world’ data to develop aggregated datasets. These could be used to promote multi-morbid clinical research, which is vital in furthering clinical practices associated with pain medicine.

## Limitations

Unified approaches of conducting DM application assessments were lacking across all 3 categories identified and reported within the scope of this study. As a result, the pooled analysis conducted limits the generalizability of the findings. It is evident that the lack of validation in digital applications is another rate-limiting factor in furthering the use of these among clinical populations.

## Conclusion

The pain medicine ecosystem has a plethora of research studies, although those in population research, prevention, clinical trials, and education, as well as training, need to evolve if improvements are to be made clinically. This could integrate evolving DM concepts, including AI applications, that could improve patient-reported outcomes. It is, therefore, important to conduct further well-designed digital clinical trials.

Another concern based on evidence ascertained in this study is the minimal use of clinical trials to test DM applications; therefore, the efficacy and efficiency of these, as well as the generalizability to a wider population, remain limited. Pragmatic and novel methods of conducting clinical trials would be beneficial in providing credible evidence before these DM applications are used within clinical practice. Alternatives such as simulation studies using *real-world* environments could be used to test novel DM applications, given the complexities around conducting pain research. Similarly, it may be beneficial for patients to gain access to DM applications more quickly, especially those managing chronic pain. Therefore, a paradox of “*no evidence, no implementation vs. no implementation, no evidence*” is a challenge to clinicians, patients, policymakers, and clinical researchers alike. Using simulation methods, where possible, could provide an alternative method to overcome this paradox, although there may be limitations that would need considering as it not always feasible to design precise simulations or perform competency validation. The proliferation of digital technologies would provide the leverage to optimise global care by way of mobile platforms, to open better avenues, and to measure outcome data from wearable devices. These applications use real-world data that could benefit patients and clinicians alike. Thus, the use of DM in pain medicine could promote a myriad of benefits.

## Supporting information

Characteristic table

## Data Availability

All data produced in the present study are available upon reasonable request to the authors

## Acknowledgments

This paper is part of the multifaceted ELEMI project that is sponsored by Southern Health NHS Foundation Trust and in collaboration with the University of Liverpool, University College London, University College London NHS Foundation Trust, Liverpool Women’s Hospital, University of Southampton, Robinson Institute-University of Adelaide, Ramaiah Memorial Hospital (India), University of Geneva and Manchester University NHS Foundation Trust.

This paper is also part of the POP project focusing on Chronic Pain that is sponsored by University College London NHS Foundation Trust and in collaboration with University of Oxford and University of Southampton.

## Author Contributions

GD conceptualised the logic model of this paper and developed the systematic methodology. GD and AS wrote the first draft of the manuscript. The systematic methodology was critically appraised by AS and GD initially. RE, GD and AS extracted the data. This data was reviewed by GD and AS. GD, JS and YZ developed the statistical analysis plan. JS, YZ, GD, TF and AS conducted the full analysis. The full paper was critically appraised by all authors. All authors approved the final version of the manuscript.

## Data Availability Statement

All data used within this study has been publicly available. The authors will consider sharing the dataset gathered upon request.

## References

1. Edwards RR, Dworkin RH, Turk DC, Angst MS, Dionne R, Freeman R, et al. Patient phenotyping in clinical trials of chronic pain treatments: IMMPACT recommendations. Pain. 2016;157(9):1851–1871. doi:10.1097/j.pain.0000000000000602

2. Rosenblum A, Marsch LA, Joseph H, Portenoy RK. Opioids and the treatment of chronic pain: controversies, current status, and future directions. Exp Clin Psychopharmacol. 2008;16(5):405–416. doi:10.1037/a0013628

3. Broderick JE, Schwartz JE, Vikingstad G, Pribbernow M, Grossman S, Stone AA. The accuracy of pain and fatigue items across different reporting periods. Pain. 2008;139(1):146–157. doi:10.1016/j.pain.2008.03.024

4. Bolger N, Laurenceau J. Intensive longitudinal methods: An introduction to diary and experience sampling research. Guilford Press; 2013.

5. Fillingim RB, Loeser JD, Baron R, Edwards RR. Assessment of Chronic Pain: Domains, Methods, and Mechanisms. J Pain. 2016;17(9 Suppl):T10–T20. doi:10.1016/j.jpain.2015.08.010

6. Mun CJ, Suk HW, Davis MC, Karoly P, Finan P, Tennen H, et al. Investigating intraindividual pain variability: methods, applications, issues, and directions. Pain. 2019;160(11):2415–2429. doi:10.1097/j.pain.0000000000001626

7. World Health Organization. Monitoring and Evaluating Digital Health Interventions: a practical guide to conducting research and assessment. http://apps.who.int/iris/bitstream/handle/10665/252183/9789241511766-eng.pdf;jsessionid=9630003E91620D111417E2CE52AF8075?sequence=1. Accessed 23 May, 2021.

8. Medicines & Healthcare products Regulatory Agency. Custom-made devices in Great Britain. https://www.gov.uk/government/publications/custom-made-medical-devices/custom-made-devices-in-great-britain. Accessed 23 May, 2021.

9. U.S. Food and Drug Administration. Factors to Consider Regarding Benefit Risk in Medical Device Product Availability, Compliance, and Enforcement Decisions: Guidance for Industry and Food and Drug Administration Staff. https://www.fda.gov/files/medical%20devices/published/Factors-to-Consider-Regarding-Benefit-Risk-in-Medical-Device-Product-Availability--Compliance--and-Enforcement-Decisions---Guidance-for-Industry-and-Food-and-Drug-Administration-Staff.pdf. Accessed 23 May, 2021.

10. National Institute for Health and Care Excellence. Evidence standards framework for digital health technologies. https://www.nice.org.uk/Media/Default/About/what-we-do/our-programmes/evidence-standards-framework/digital-evidence-standards-framework.pdf. Accessed 23 May, 2021.

11. Gentili C, Zetterqvist V, Rickardsson J, Holmström L, Simons LE, Wicksell RK. ACTsmart: Guided Smartphone-Delivered Acceptance and Commitment Therapy for Chronic Pain-A Pilot Trial. Pain Med. 2021;22(2):315–328. doi:10.1093/pm/pnaa360

12. Bostrøm K, Børøsund E, Varsi C, Eide H, Nordang EF, Schreurs KM, et al. Digital Self-Management in Support of Patients Living With Chronic Pain: Feasibility Pilot Study. JMIR Form Res. 2020;4(10):e23893. doi:10.2196/23893

13. Greenberg J, Popok PJ, Lin A, Kulich RJ, James P, Macklin EA, et al. A Mind-Body Physical Activity Program for Chronic Pain With or Without a Digital Monitoring Device: Proof-of-Concept Feasibility Randomized Controlled Trial. JMIR Form Res. 2020;4(6):e18703. doi:10.2196/18703

14. Jonassaint CR, Rao N, Sciuto A, Switzer GE, De Castro L, Kato GJ, et al. Abstract Animations for the Communication and Assessment of Pain in Adults: Cross-Sectional Feasibility Study. J Med Internet Res. 2018;20(8):e10056. doi:10.2196/10056

15. van der Heijden AA, Abramoff MD, Verbraak F, van Hecke MV, Liem A, Nijpels G. Validation of automated screening for referable diabetic retinopathy with the IDx-DR device in the Hoorn Diabetes Care System. Acta Ophthalmol. 2018;96(1):63–68. doi:10.1111/aos.13613

16. Bossen D, Veenhof C, Van Beek KE, Spreeuwenberg PM, Dekker J, De Bakker DH. Effectiveness of a web-based physical activity intervention in patients with knee and/or hip osteoarthritis: randomized controlled trial. J Med Internet Res. 2013;15(11):e257. doi:10.2196/jmir.2662

17. Hedman-Lagerlöf M, Hedman-Lagerlöf E, Axelsson E, Ljótsson B, Engelbrektsson J, Hultkrantz S, et al. Internet-Delivered Exposure Therapy for Fibromyalgia: A Randomized Controlled Trial. Clin J Pain. 2018;34(6):532–542. doi:10.1097/AJP.0000000000000566

18. Krein SL, Kadri R, Hughes M, Kerr EA, Piette JD, Holleman R, et al. Pedometer-based internet-mediated intervention for adults with chronic low back pain: randomized controlled trial. J Med Internet Res. 2013;15(8):e181. doi:10.2196/jmir.2605

19. Rini C, Porter LS, Somers TJ, McKee DC, DeVellis RF, Smith M, et al. Automated Internet-based pain coping skills training to manage osteoarthritis pain: a randomized controlled trial. Pain. 2015;156(5):837–848. doi:10.1097/j.pain.0000000000000121

20. Williams DA, Kuper D, Segar M, Mohan N, Sheth M, Clauw DJ. Internet-enhanced management of fibromyalgia: a randomized controlled trial. Pain. 2010;151(3):694–702. doi:10.1016/j.pain.2010.08.034

21. Wilson M, Roll JM, Corbett C, Barbosa-Leiker C. Empowering Patients with Persistent Pain Using an Internet-based Self-Management Program. Pain Manag Nurs. 2015;16(4):503–514. doi:10.1016/j.pmn.2014.09.009

22. Raj SX, Brunelli C, Klepstad P, Kaasa S. COMBAT study - Computer based assessment and treatment - A clinical trial evaluating impact of a computerized clinical decision support tool on pain in cancer patients. Scand J Pain. 2017;17:99–106. doi:10.1016/j.sjpain.2017.07.016

23. Guillory J, Chang P, Henderson Jr CR, Shengelia R, Lama S, Warmington M, et al. Piloting a Text Message-based Social Support Intervention for Patients With Chronic Pain: Establishing Feasibility and Preliminary Efficacy. Clin J Pain. 2015;31(6):548–556. doi:10.1097/AJP.0000000000000193

24. Berman RL, Iris MA, Bode R, Drengenberg C. The effectiveness of an online mind-body intervention for older adults with chronic pain. J Pain. 2009;10(1):68–79. doi:10.1016/j.jpain.2008.07.006

25. Carpenter KM, Stoner SA, Mundt JM, Stoelb B. An online self-help CBT intervention for chronic lower back pain. Clin J Pain. 2012;28(1):14–22. doi:10.1097/AJP.0b013e31822363db

26. Menga G, Ing S, Khan O, Dupre B, Dornelles AC, Alarakhia A, et al. Fibromyalgia: can online cognitive behavioral therapy help? Ochsner J. 2014;14(3):343–349. http://www.ochsnerjournal.org/content/14/3/343. Accessed 15 February, 2022.

27. O’moore KA, Newby JM, Andrews G, Hunter DJ, Bennell K, Smith J, et al. Internet Cognitive-Behavioral Therapy for Depression in Older Adults with Knee Osteoarthritis: A Randomized Controlled Trial. Arthritis Care Res (Hoboken). 2018;70(1):61–70. doi:10.1002/acr.23257

28. Minen MT, Adhikari S, Seng EK, Jinich S, Powers SW, Lipton RB. Smartphone-based migraine behavioral therapy: a single-arm study with assessment of mental health predictors. NPJ Digit Med. 2019;2:46. doi:10.1038/s41746-019-0116-y

29. Toelle TR, Utpadel-Fischler DA, Haas KK, Priebe JA. App-based multidisciplinary back pain treatment versus combined physiotherapy plus online education: a randomized controlled trial. NPJ Digit Med. 2019;2:34. doi:10.1038/s41746-019-0109-x

30. Blödt S, Pach D, Eisenhart-Rothe SV, Lotz F, Roll S, Icke K, et al. Effectiveness of app-based self-acupressure for women with menstrual pain compared to usual care: a randomized pragmatic trial. Am J Obstet Gynecol. 2018;218(2):227.e1-227.e9. doi:10.1016/j.ajog.2017.11.570

31. Irvine AB, Russell H, Manocchia M, Mino DE, Glassen TC, Morgan R. Mobile-Web app to self-manage low back pain: randomized controlled trial. J Med Internet Res. 2015;17(1):e3130. doi:10.2196/jmir.3130

32. Schatz J, Schlenz AM, McClellan CB, Puffer ES, Hardy S, Pfeiffer M, et al. Changes in coping, pain, and activity after cognitive-behavioral training: a randomized clinical trial for pediatric sickle cell disease using smartphones. Clin J Pain. 2015;31(6):536–547. doi:10.1097/AJP.0000000000000183

33. Skrepnik N, Spitzer A, Altman R, Hoekstra J, Stewart J, Toselli R. Assessing the Impact of a Novel Smartphone Application Compared With Standard Follow-Up on Mobility of Patients With Knee Osteoarthritis Following Treatment With Hylan G-F 20: A Randomized Controlled Trial. JMIR Mhealth Uhealth. 2017;5(5):e64. doi:10.2196/mhealth.7179

34. Sun Y, Jiang F, Gu JJ, Wang YK, Hua H, Li J, et al. Development and Testing of an Intelligent Pain Management System (IPMS) on Mobile Phones Through a Randomized Trial Among Chinese Cancer Patients: A New Approach in Cancer Pain Management. JMIR Mhealth Uhealth. 2017;5(7):e108. doi:10.2196/mhealth.7178

35. Guétin S, Brun L, Deniaud M, Clerc JM, Thayer JF, Koenig J. Smartphone-based Music Listening to Reduce Pain and Anxiety Before Coronarography: A Focus on Sex Differences. Altern Ther Health Med. 2016;22(4):60–63.

36. Jamison RN, Jurcik DC, Edwards RR, Huang CC, Ross EL. A Pilot Comparison of a Smartphone App With or Without 2-Way Messaging Among Chronic Pain Patients: Who Benefits From a Pain App? Clin J Pain. 2017;33(8):676–686. doi:10.1097/AJP.0000000000000455

37. Jibb LA, Stevens BJ, Nathan PC, Seto E, Cafazzo JA, Johnston DL, et al. Implementation and preliminary effectiveness of a real-time pain management smartphone app for adolescents with cancer: A multicenter pilot clinical study. Pediatr Blood Cancer. 2017;64(10):e26554. doi:10.1002/pbc.26554

38. Lee M, Lee SH, Kim T, Yoo HJ, Kim SH, Suh DW. Feasibility of a Smartphone-Based Exercise Program for Office Workers With Neck Pain: An Individualized Approach Using a Self-Classification Algorithm. Arch Phys Med Rehabil. 2017;98(1):80–87. doi:10.1016/j.apmr.2016.09.002

39. Oldenmenger WH, Witkamp FE, Bromberg JE, Jongen JL, Lieverse PJ, Huygen FJ, et al. To be in pain (or not): a computer enables outpatients to inform their physician. Ann Oncol. 2016;27(9):1776–1781. doi:10.1093/annonc/mdw250

40. Huber S, Priebe JA, Baumann KM, Plidschun A, Schiessl C, Tölle TR. Treatment of Low Back Pain with a Digital Multidisciplinary Pain Treatment App: Short-Term Results. JMIR Rehabil Assist Technol. 2017;4(2):e11. doi:10.2196/rehab.9032

41. Calner T, Nordin C, Eriksson MK, Nyberg L, Gard G, Michaelson P. Effects of a self-guided, web-based activity programme for patients with persistent musculoskeletal pain in primary healthcare: A randomized controlled trial. Eur J Pain. 2017;21(6):1110–1120. doi:10.1002/ejp.1012

42. Chiauzzi E, Pujol LA, Wood M, Bond K, Black R, Yiu E, et al. painACTION-back pain: a self-management website for people with chronic back pain. Pain Med. 2010;11(7):1044–1058. doi:j.1526-4637.2010.00879.x

43. Davis MC, Zautra AJ. An online mindfulness intervention targeting socioemotional regulation in fibromyalgia: results of a randomized controlled trial. Ann Behav Med. 2013;46(3):273–284. doi:10.1007/s12160-013-9513-7

44. Dowd H, Hogan MJ, McGuire BE, Davis MC, Sarma KM, Fish RA, et al. Comparison of an Online Mindfulness-based Cognitive Therapy Intervention With Online Pain Management Psychoeducation: A Randomized Controlled Study. Clin J Pain. 2015;31(6):517–527. doi:10.1097/AJP.0000000000000201

45. Lin J, Wurst R, Paganini S, Hohberg V, Kinkel S, Göhner W, et al. A group-and smartphone-based psychological intervention to increase and maintain physical activity in patients with musculoskeletal conditions: study protocol for a randomized controlled trial (“MoVo-App”). Trials. 2020;21(1):502. doi:10.1186/s13063-020-04438-4

46. Nordin CA, Michaelson P, Gard G, Eriksson MK. Effects of the Web Behavior Change Program for Activity and Multimodal Pain Rehabilitation: Randomized Controlled Trial. J Med Internet Res. 2016;18(10):e265. doi:10.2196/jmir.5634

47. Ruehlman LS, Karoly P, Enders C. A randomized controlled evaluation of an online chronic pain self management program. Pain. 2012;153(2):319–330. doi:10.1016/j.pain.2011.10.025

48. Ström L, Pettersson R, Andersson G. A controlled trial of self-help treatment of recurrent headache conducted via the Internet. J Consult Clin Psychol. 2000;68(4):722–727. doi:10.1037/0022-006x.68.4.722

49. Anderson KO, Mendoza TR, Payne R, Valero V, Palos GR, Nazario A, et al. Pain education for underserved minority cancer patients: a randomized controlled trial. J Clin Oncol. 2004;22(24):4918–4925. doi:10.1200/JCO.2004.06.115

50. Lovell MR, Forder PM, Stockler MR, Butow P, Briganti EM, Chye R, et al. A randomized controlled trial of a standardized educational intervention for patients with cancer pain. J Pain Symptom Manage. 2010;40(1):49–59. doi:10.1016/j.jpainsymman.2009.12.013

51. Guétin S, Brun L, Mériadec C, Camus E, Deniaud M, Thayer JF, et al. A Smartphone-Based Music Intervention to Reduce Pain and Anxiety in Women before or during labor. EuJIM. 2018;21:24–26. doi:10.1016/j.eujim.2018.06.001

52. Oldenmenger WH, Baan MAG, van der Rijt CCD. Development and feasibility of a web application to monitor patients’ cancer-related pain. Support Care Cancer. 2018;26(2):635–642. doi:10.1007/s00520-017-3877-3

53. Kamdar M, Centi AJ, Agboola S, Fischer N, Rinaldi S, Strand JJ, et al. A randomized controlled trial of a novel artificial intelligence-based smartphone application to optimize the management of cancer-related pain. J Clin Onc. 2019;37:11514. doi:10.1200/JCO.2019.37.15_suppl.11514

54. Solem IK, Varsi C, Eide H, Kristjansdottir OB, Mirkovic J, Børøsund E, et al. Patients’ Needs and Requirements for eHealth Pain Management Interventions: Qualitative Study. J Med Internet Res. 2019;21(4):e13205. doi:10.2196/13205

55. Witzeman K, Flores OA, Renzelli-Cain RI, Worly B, Moulder JK, Carrillo JF, et al. Patient-Physician Interactions Regarding Dyspareunia with Endometriosis: Online Survey Results. J Pain Res. 2020;13:1579–1589. doi:10.2147/JPR.S248887

56. McConnell MV, Shcherbina A, Pavlovic A, Homburger JR, Goldfeder RL, Waggot D, et al. Feasibility of Obtaining Measures of Lifestyle From a Smartphone App: The MyHeart Counts Cardiovascular Health Study. JAMA Cardiol. 2017;2(1):67–76. doi:10.1001/jamacardio.2016.4395

57. Urteaga I, McKillop M, Elhadad N. Learning endometriosis phenotypes from patient-generated data. NPJ Digit Med. 2020;3:88. doi:10.1038/s41746-020-0292-9

58. Jacobson NC, Summers B, Wilhelm S. Digital Biomarkers of Social Anxiety Severity: Digital Phenotyping Using Passive Smartphone Sensors. J Med Internet Res. 2020;22(5):e16875. doi:10.2196/16875

59. Inan OT, Tenaerts P, Prindiville SA, Reynolds HR, Dizon DS, Cooper-Arnold K, et al. Digitizing clinical trials. NPJ Digit Med. 2020;3(101):1–7. doi:10.1038/s41746-020-0302-y

60. Guo C, Ashrafian H, Ghafur S, Fontana G, Gardner C, Prime M. Challenges for the evaluation of digital health solutions-A call for innovative evidence generation approaches. NPJ Digit Med. 2020;3:110. doi:10.1038/s41746-020-00314-2

