## Supplementary material for "A Systematic Review and Meta-Analysis of Digital Application use in Clinical Research in Pain Medicine": Characteristic table

### Table 1. Key clinical features and classification of pain

| Categories of pain | Summary | Interventions-Drug | Interventions-Medical devices |
|---|---|---|---|
| Nociceptive pain | Somatic or visceral pain arises from actual or threatened damage to non-neural tissue and is due to the activation of nociceptors. | Physical therapy, Medications, Injection, Treatment of the underlying cause NSAID, | Injection, Treatment of the underlying cause |
| Inflammatory pain | Cardinal feature of an inflammatory state of hypersensitivity where innocuous stimuli induce pain. This is regulated by prostaglandin receptors EPI1, EP2, EP3 and EP4. | Anti-inflammatory medicine such as Aspirin, Ibuprofen, Diclofenac, Flurbiprofen, Mefanamic acid, Nabumetone, Piroxicam and Naproxen | Laser therapy systems, electrotherapy device, non-invasive heat therapy, infrared light therapy, pulsed electromagnetic field therapy and electrode induced pain blocker. |
| Dysfunctional/Functional Pain | Pain without obvious organic cause. | Physical Therapy, Cognitive Behavioural Therapy (CBT) | TENS, External Neuromodulation |
| Neuropathic pain | Pain caused by a lesion or disease of the somatosensory nervous system. | Anti-neuropathic medication | Neuromodulation, Nerve blocks |

### Table 2. Description of Regulatory Guidelines by Organisation

| Organisation/Regulator | Data published | Guidelines/Regulations | Description |
|---|---|---|---|
| MHRA (Medicines and Healthcare Products Regulatory Agency) | 2020 | Medical Device Regulations (MDR) | The medical device directive was amended to the MDR to accommodate artificial intelligence applications that provide medical advice or act as a clinician aid. In addition, the UK conformity assessed (UKCA) marking that is equivalent to CE marking. The UKCA marking is a standard required for goods placed on the market in Great Britain |
| Code of conduct for data-driven health and care technology | 2019 | A policy document with 10 principles to evaluate digital health solutions | There is an associated code of conduct within the United Kingdom |
| NICE (National Institute of Health and Care Excellence) evidence standards framework | 2019 | The evidence tiers are cumulative therefore, the framework aims to assess digital health technologies (DHT) from a specific risk and best practice perspective. | The framework demonstrates the standards required for the evidence developed or available about digital health technologies that may or may not fall within the remit of medical devices, including those associated with artificial intelligence. This framework includes evidence associated with effectiveness specific to the intended purposes and user. This also includes evidence associated with the economic impact affiliated with financial risk. |
| FDA (Food and Drug Administration) | 2016 | Benefit-risk framework for medical devices | A generic framework to evaluate medical devices risks and benefits. For example, this framework can assess dimensions such as risk serenity and the likelihood of risk as well as false positive or false-negative results. |
| WHO (World Health Organisation) | 2016 | WHO monitoring and evaluating digital health interventions | A generic framework to evaluate and validate digital solutions throughout the life-cycle of the innovation. |
| IMDRF (International Medical Device Regulation Forum) | 2017 | Multiple definitions are included and developed by the International Medical Device Regulators to manage these across multiple countries | The scope of this framework is a foundational approach to address unique challenges which include common vocabulary to identify specific information to support healthcare decision making |

| | | | |
|---|---|---|---|
| | | | and/or healthcare conditions with core functions |
| MEDDEV 2.1 (Medical Device Regulations) | 2016 | Guidelines on the quantification and classification of standalone software with a medical purpose and are associated within the regulatory framework of medical devices. These aspects are part of the European Union | These guidelines describe any medical tool when they do not fall within the remit of software, or in vitro diagnostic devices or a medical device |
| FDA (Food and Drug Administration) pre-certification program | 2019 | This is a software pre-certification program developed as a pilot | This program includes a test plan designed to evaluate the Excellence Appraisal and Streamlined Review composites that would formulate quality assurance for the safety and effectiveness of the software prior to implementation clinically. |
| FDA (Food and Drug Administration)-SaMD (Proposed regulatory framework for modifications to artificial intelligence/machine learning such as software as a medical device | 2019 | SaMD demonstrates a risk categorisation framework inclusive of a risk-based approach to categorise intended use | This framework assesses 2 key descriptions of intended use; state of the healthcare situation or clinical condition and the patient population it is intended for in terms of risk (critical; serious, or non-serious healthcare situation/condition; and the information provided by the SaMD to formulate the decision for the intended user to diagnose or treat or clinically manage the condition. |
| FDA (Food and Drug Administration) Medical mobile application guidance | 2018 | The purpose of this guidance is for mobile platform as defined as a commercial computer platform with or without wireless connectivity that could be hand handled such as a platform downloadable to a smart phone or tablet. | This guidelines document is relevant to assess mobile applications where it is a software that is tailored to a mobile platform. This also includes device software functions defined under section 201(h) of the FD&C Act which could be conformed as an accessory to an existing regulated medical device or to transform a mobile platform into a regulated medical device. |

**Table 3. Summary table of current uses of digital applications within clinical medicine**

| Benefits | Summary | Application examples |
|---|---|---|
| Improved access | Use of smart-devices intra-connected linking various streams together | Apple watch gathering step-count data as part of their exercise app |
| Effective data collection | Self-reported data gathering is useful in particular for healthcare professionals | Healthy; The self-care App which acts as diary cards for anyone with a health condition |
| Efficient data processing | Processing big data is useful to better understand the disease profile and potential treatment avenues | NHS Weight Loss Plan App acts as a support tool to anyone attempting to lose weight |
| Early diagnosis | Using data to make an early diagnosis through symptom tracking and/or identification of pattern inferences within gathered data is useful for healthcare systems, patients and clinicians | Sleepio App is used currently to support those with sleep-related issues. The app allows data to be gathered to personalise strategies to help improve sleep quality |
| Personalised treatment | Using data with patient preferences would aid development of personalised treatment | Various applications are currently available within a research context and are yet to be implemented clinically |

**Table 4 Characteristics of the systematically included studies**

| Author | Diagnosis/Treatment method | Digital application and method of application delivery | Study type | Sample size | Country | Exposure |
|---|---|---|---|---|---|---|
| Bossen et al (2013) | Intervention | Web-based intervention | RCT | 199 | Netherlands | osteoarthritis pain |
| Hedman-Lagerlöf, et al (2018) | Intervention | Web-based intervention | RCT | 140 | Sweden | fibromyalgia |
| Krein et al (2013) | Intervention | Web-based intervention | RCT | 229 | USA | chronic low back pain |
| Rini et al (2015) | Intervention | Web-based intervention | RCT | 113 | USA | osteoarthritis pain |
| Williams et al (2010) | Intervention | Web-based intervention | RCT | 118 | USA | fibromyalgia |
| Wilson et al (2015) | Intervention | Web-based intervention | RCT | 92 | USA | chronic non-cancer pain |
| Raj et al (2017) | Intervention | Web-based intervention | RCT | 214 | Norway | cancer-related pain |
| Guillory et al (2015) | Chatbots | Text message and mobile app | RCT-Feasibility | 68 | USA | chronic non-cancer pain |
| Berman et al (2009) | Chatbots | Web-based intervention | RCT | 78 | USA | chronic pain |
| Carpenter et al (2012) | Chatbots-Cognitive behavioral therapy with chapters | Web-based intervention | RCT-Pilot | 141 | USA | chronic low back pain |
| Menga et al (2014) | Chatbots-Cognitive behavioral therapy with chapters | Web-based intervention | RCT | 44 | USA | Fibromyalgia |
| O'moore et al (2018) | Chatbots-Cognitive behavioral therapy with chapters | Web-based intervention | RCT | 69 | USA | osteoarthritis pain |
| Gentili et al (2020) | Mobile app based acceptance therapy | Mobile based intervention | RCT-pilot | 31 | Sweden | chronic pain |
| Minen et al (2019) | Mobile app based behavioral therapy | Mobile based intervention | Cross-sectional - Feasibility | 51 | USA | migraine |
| Toelle et al (2019) | Mobile app based therapy | Mobile based intervention | RCT | 94 | Germany | Chronic non-specific low back pain |
| Blödt et al (2018) | Mobile app based self-acupressure | Mobile based intervention | RCT-Pragmatic | 221 | Germany | menstrual pain |
| Irvine et al (2015) | Mobile app based self-management | Mobile based intervention | RCT | 597 | USA | chronic low back pain |
| Schatz et al (2015) | Mobile app based coping, pain and activity | Mobile based intervention | RCT | 46 | USA | chronic pain for paediatric sickle cell |
| Nebojsa et al (2017) | Mobile app and an wearable activity monitor | Mobile based intervention | RCT | 211 | USA | osteoarthritis pain |
| Sun et al (2017) | Mobile app for pain management | Mobile based intervention | RCT | 46 | China | cancer related pain |
| Guétin et al (2016) | Mobile app delivering music therapy for pain | Mobile based intervention | RCT | 106 | France | chronic pain |

| | | | | | | |
|---|---|---|---|---|---|---|
| Jamison et al (2017) | Mobile app based daily assessment and treatment | Mobile based intervention | RCT-pilot | 90 | USA | chronic pain |
| Jibb et al (2017) | Mobile apps | Mobile based intervention | RCT-pragmatic | 40 | Canada | cancer-related chronic pain among the adolescent |
| Lee et al (2017) | Mobile app based exercise program | Mobile based intervention | Cross-section single group repeated measure | 23 | Korea | neck pain |
| Oldenmenger et al (2016) | Mobile apps | Web-based intervention | quantitative | 48 | Netherlands | cancer-related pain |
| Huber et al (2017) | Mobile app and EHR | Mobile based intervention | Retrospective RCT | 180 | Germany | chronic low back pain |
| Calner et al (2017) | Intervention | Web-based intervention | RCT | 109 | Sweden | musculoskeletal pain |
| Chiauzzi et al (2010) | Intervention-self management | Web-based intervention | RCT | 199 | USA | chronic pain |
| Davis et al (2013) | Intervention of mindfulness | Web-based intervention | RCT | 79 | USA | fibromyalgia |
| Dowd et al (2015) | Online mindfulness based cognitive therapy intervention | Web-based intervention | RCT | 124 | Ireland | chronic pain |
| Lin et al (2020) | Mobile apps | Web-based intervention | RCT | 302 | Germany | multimodal pain |
| Nordin et al (2016) | Intervention for web behaviour change | Web-based intervention | RCT | 109 | Sweden | Multimodal pain |
| Ruehlmana et al (2012) | Intervention-self management | Web-based intervention | RCT | 305 | USA | chronic pain |
| Ström et al (2000) | Intervention-self management | Web-based intervention | RCT | 45 | Sweden | recurrent headache |
| Anderson et al (2004) | Intervention-video and booklet | Web-based intervention | RCT | 97 | USA | Cancer related pain |
| Lovell et al (2010) | Intervention-video and booklet | Web-based intervention | RCT | 217 | Australia | Cancer related pain |
| Guétin et al (2018) | Smart phone based intervention | Mobile based intervention | RCT | 62 | France | chronic painful conditions |
| Oldenmenger et al (2018) | Intervention-internet applications | Web-based intervention | cohort study | 84 | Netherlands | Cancer related pain |

**EHR: Electronic Health Records RCT-Randomised clinical trial*

### Table 5 Studies included within the meta-analysis

| Study ID | Author | Digital applications | Study type | Sample size | Country | Exposure | P-value |
|---|---|---|---|---|---|---|---|
| 1 | Bossen et al (2013) | web-application | RCT | 199 | Netherlands | osteoarthritis pain | 0.33(pain intensity) 0.09(depression) 0.007(anxiety) |
| 2 | Hedman-Lagerlöf et al (2018) | web-application | RCT | 140 | Sweden | fibromyalgia | <0.001(depression) <0.001(anxi |

| | | | | | | | |
|---|---|---|---|---|---|---|---|
| | | | | | | | ety)<br><0.001(fatigue) |
| 3 | Rini et al (2015) | web-application | RCT | 113 | USA | osteoarthritis pain | Not provided |
| 4 | Williams et al (2010) | web-application | RCT | 118 | USA | fibromyalgia | Not provided |
| 5 | Wilson et al (2015) | web-application | RCT | 92 | USA | chronic noncancer pain | 0.22(pain intensity)<br>0.25(depression) |
| 6 | Raj et al (2017) | web-application | RCT | 214 | Norway | cancer-related pain | Not provided |
| 7 | Berman et al (2019) | chatbots | RCT | 78 | USA | chronic pain | Not provided |
| 8 | Menga et al (2014) | chatbots | RCT | 44 | USA | Fibromyalgia | 0.005 (severity of fibromyalgia) |
| 9 | O'moore et al (2018) | chatbots | RCT | 69 | Australia | osteoarthritis pain | Not provided |
| 10 | Gentili et al (2020) | Mobile apps | RCT | 94 | Germany | chronic low back pain | 0.021(pain intensity) |
| 11 | Blödt et al (2018) | Mobile apps | RCT | 221 | Germany | menstrual pain | 0.026(pain intensity) |
| 12 | Schatz et al (2015) | Mobile apps | RCT | 46 | USA | chronic pain | 0.1(negative affect) |
| 13 | Sun et al (2017) | Mobile apps | RCT | 46 | China | cancer-related pain | <0.01(pain intensity) |
| 14 | Calner et al (2017) | Mobile apps | RCT | 109 | USA | musculoskeletal pain | 0.37(intensity) |
| 15 | Chiauzzi et al (2010) | Mobile apps | RCT | 199 | USA | chronic pain | Not provided |
| 16 | Dowd et al (2015) | Mobile apps | RCT | 124 | Ireland | chronic pain | Not provided |
| 17 | Lin et al (2020) | Mobile apps | RCT | 302 | Germany | multimodal pain | 0.01(pain intensity)<br><0.01(depression)<br>0.44(anxiety)<br><0.01(pain interference) |
| 18 | Ruehlmana et al (2012) | Mobile apps | RCT | 305 | USA | chronic pain | 0.2(pain intensity)<br>0.06(depression)<br>0.15(anxiety)<br>0.3(pain interference) |
| 19 | Ström et al (2000) | Mobile apps | RCT | 45 | Sweden | recurrent headache | Not provided |
| 20 | Anderson et al (2004) | web-application | RCT | 84 | Netherlands | Cancer related pain | Not provided |

**Table 6 Risk of bias, according to the revised risk-of-bias tool for randomised trials (RoB 2.0)**

| Author | Randomization Process | Deviations from the intended interventions | Missing Outcome Data | Measurement of the Outcome | Selection of the reported result | Overall |
|---|---|---|---|---|---|---|
| Bossen et al (2013) | some concerns* | low risk | low risk | low risk | low risk | some concerns |
| Hedman-Lagerlöf, et al (2018) | low risk | low risk | low risk | low risk | low risk | low risk |
| Krein et al (2013) | low risk | low risk | low risk | low risk | low risk | low risk |
| Rini et al (2015) | low risk | low risk | low risk | low risk | low risk | low risk |
| Williams et al (2010) | low risk | low risk | low risk | low risk | low risk | low risk |
| Wilson et al (2015) | low risk | low risk | low risk | low risk | low risk | low risk |
| Raj et al (2017) | low risk | low risk | low risk | low risk | low risk | low risk |
| Guillory et al (2015) | low risk | low risk | low risk | low risk | low risk | low risk |
| Berman et al (2009) | low risk | low risk | low risk | low risk | low risk | low risk |

| | | | | | | |
|---|---|---|---|---|---|---|
| Carpenter et al (2012) | low risk | low risk | low risk | low risk | low risk | low risk |
| Menga et al (2014) | low risk | low risk | low risk | low risk | low risk | low risk |
| O'moore et al (2018) | low risk | low risk | low risk | low risk | low risk | low risk |
| Gentili et al (2020) | low risk | low risk | low risk | low risk | low risk | low risk |
| Minen et al (2019) | high risk** | low risk | low risk | low risk | low risk | high risk |
| Toelle et al (2019) | some concerns*** | low risk | low risk | low risk | low risk | some concerns |
| Blödt et al (2018) | low risk | low risk | low risk | low risk | low risk | low risk |
| Irvine et al (2015) | low risk | low risk | low risk | low risk | low risk | low risk |
| Schatz et al (2015) | low risk | low risk | low risk | low risk | low risk | low risk |
| Nebojsa et al (2017) | low risk | low risk | low risk | low risk | low risk | low risk |
| Sun et al (2017) | low risk | low risk | low risk | low risk | low risk | low risk |
| Guétin et al (2016) | low risk | low risk | low risk | low risk | low risk | low risk |
| Jamison et al (2017) | low risk | low risk | low risk | low risk | low risk | low risk |
| Jibb et al (2017) | low risk | low risk | low risk | low risk | low risk | low risk |
| Lee et al (2017) | high risk** | low risk | low risk | low risk | low risk | high risk |
| Oldenmenger et al (2016) | low risk | low risk | low risk | low risk | low risk | low risk |
| Huber et al (2017) | low risk | low risk | low risk | low risk | low risk | low risk |
| Calner et al (2017) | low risk | low risk | low risk | low risk | low risk | low risk |
| Chiauzzi et al (2010) | low risk | low risk | low risk | low risk | low risk | low risk |
| Davis et al (2013) | low risk | low risk | low risk | low risk | low risk | low risk |
| Dowd et al (2015) | low risk | low risk | low risk | low risk | low risk | low risk |
| Lin et al (2020) | low risk | low risk | low risk | low risk | low risk | low risk |
| Nordin et al (2016) | low risk | low risk | low risk | low risk | low risk | low risk |
| Ruehlmana et al (2012) | low risk | low risk | low risk | low risk | low risk | low risk |
| Ström et al (2000) | low risk | low risk | low risk | low risk | low risk | low risk |
| Anderson et al (2004) | low risk | low risk | low risk | low risk | low risk | low risk |
| Lovell et al (2010) | low risk | low risk | low risk | low risk | low risk | low risk |
| Guétin et al (2018) | low risk | low risk | low risk | low risk | low risk | low risk |
| Oldenmenger et al (2018) | high risk** | low risk | low risk | low risk | low risk | high risk |

* Some concerns due to missing information regarding the allocation concealment.**High risk because of lack of randomisation.
*** Some concerns due to deviation from the protocol resulting in a 53:48 distribution of participants.
